# Nanopore sequencing for *Mycobacterium tuberculosis* drug susceptibility testing and outbreak investigation

**DOI:** 10.1101/2022.03.04.22271870

**Authors:** Michael B. Hall, Marie Sylvianne Rabodoarivelo, Anastasia Koch, Anzaan Dippenaar, Sophie George, Melanie Grobbelaar, Robin Warren, Timothy Walker, Helen Cox, Sebastien Gagneux, Derrick Crook, Tim Peto, Niaina Rakotosamimanana, Simon Grandjean Lapierre, Zamin Iqbal

## Abstract

**Background:** *Mycobacterium tuberculosis* whole-genome sequencing (WGS) using Illumina technology has been widely adopted for genotypic drug susceptibility testing (DST) and outbreak investigation. Oxford Nanopore Technologies is reported to have higher error rates but has not been thoroughly evaluated for these applications.

**Methods:** We analyse 151 isolates from Madagascar, South Africa and England with phenotypic DST and matched Illumina and Nanopore data. Using PacBio assemblies, we select Nanopore filters for BCFtools (software) detection of single nucleotide polymorphisms (SNPs). We compare transmission clusters identified by Nanopore and the United Kingdom Health Security Agency Illumina pipeline (COMPASS). We compare Illumina and Nanopore WGS-based DST predictions using Mykrobe (software).

**Findings:** Nanopore/BCFtools identifies SNPs with median precision/recall of 99·5/90·2% compared with 99·6/91·9% for Illumina/COMPASS. Using a threshold of 12 SNPs for putative transmission clusters, Illumina identifies 98 isolates as unrelated and 53 as belonging to 19 distinct clusters (size range 2-7). Nanopore reproduces this distribution with addition of 5 singleton isolates to distinct clusters and merging of two cluster pairs. Illumina-based clusters are also replicated using a 5 SNP threshold. Clustering accuracy is maintained using mixed Illumina/Nanopore datasets. Genotyping resistance variants is highly concordant, with 0(4) discordant SNPs (indels) across 151 isolates genotyped at >3000 (60,000) SNPs (indels).

**Interpretation:** Illumina and Nanopore sequence data provide comparable cluster-identification and DST results.

**Funding:** Academy for Medical Sciences (SGL018\110), Oxford Wellcome Institutional Strategic Support Fund (ISSF TT17 4). Swiss South Africa Joint Research Award (Swiss national science Foundation and South African national research foundation).

**Research in context:** *Evidence before this study:* Two key types of information can be obtained from laboratory testing of *M. tuberculosis* isolates to help directly guide public health interventions: drug susceptibility testing (DST) to guide therapy, and bacterial typing to enrich understanding of the epidemiology and guide interventions to mitigate transmission. DST is typically performed by the “gold standard” culture-based phenotyping method or nucleic acid amplification assays targeting specific resistance-conferring mutations. Studies over the last 7 years have shown that prediction of susceptibility profile using Illumina-technology genome sequence data is possible, and can be automated. In a key publication, the CRyPTIC consortium and UK 100,000 Genomes project evaluated the method on over 10,000 genomes including prospectively sampled isolates and showed that for first-line tuberculosis (TB) drugs (isoniazid, rifampicin, ethambutol, pyrazinamide) a pan-susceptibility profile is accurate enough to be used clinically. The genetic basis of resistance remains imperfectly understood for second-line TB drugs, in particular for new and repurposed drugs (bedaquiline, clofazimine, delamanid, linezolid). Prior work in the field of genotypic DST was heavily based on Illumina technology, which provides short (70-300 base pair) sequence reads of very high quality. Many different softwares (e.g. TBProfiler, Mykrobe, MTBseq, kvarq) have been designed for sequence analysis and genotypic DST. However, the increasingly used Nanopore sequencing platforms yield very different data with much longer sequence reads (frequently over 1kb) and higher error rates including systematic biases. To date, very limited evaluation of Nanopore-based drug susceptibility prediction has been performed using the only two compatible tools (Mykrobe (n=5 independent samples), TBProfiler (n=3 independent samples)). Molecular typing of *M. tuberculosis* allows lineage identification and detection of putative transmission clusters. In the last decade, multiple *M. tuberculosis* molecular epidemiology studies have shown how genomic information can complement traditional epidemiology in identifying person-to-person transmission clusters with a high level of resolution. Typically, the number of single nucleotide polymorphism (SNP) disagreements between genomes, or SNP distance, is calculated and single-linkage clustering is performed for genomes falling within retrospectively established transmission thresholds of either 5 or 12 SNPs. Just as with DST, these thresholds were established with Illumina sequencing data. The increased error rate in Nanopore sequencing is believed to lead to inflated SNP distances if standard genome analysis tools are used. Prior to this study it was unknown what impact on isolate-clustering this would incur.

*Added value of this study:* Full-scale adoption of genomic sequencing in tuberculosis reference laboratories has so far taken place in a limited number of settings - England, the Netherlands, and New York State - all using Illumina-based sequencing data. Building on current evidence, specific WHO technical guidance and diversification and democratisation of technology, sequencing is expected to be increasingly used in tuberculosis control globally. For the first time, our study offers 4 key deliverables intended to inform adoption of Nanopore technology as an alternative, or a complement, to Illumina. First: a systematic head-to-head comparison of Nanopore and Illumina data for *M. tuberculosis* drug susceptibility profiling and isolate clustering, including quantitative metrics for cluster precision and recall. Second: an assessment of the impact of mixed Illumina and Nanopore data on clustering which represents an increasingly common challenge. Third: an open-source software pipeline allowing research and reference laboratories to replicate our analytical approach. Fourth: a publicly available curated test set of 151 isolates, including matched Illumina and Nanopore sequence data, and (for a subset of seven isolates) high-quality PacBio assemblies, for method development and validation.

*Implications of all the available evidence:* Catalogues of drug resistance conferring mutations will keep improving, especially for new and repurposed drugs. Our data confirms that Illumina and Nanopore sequencing technologies can be used to identify those mutations equally accurately in *M. tuberculosis*. Bacterial molecular typing is constantly shown to support the understanding of disease transmission and tuberculosis control in new settings. The bioinformatics tools and filters we have developed, assessed, and made publicly available allow the use of Nanopore or mixed-technology data to appropriately cluster genetically related isolates. We provide a measure of the expected level of over-clustering associated with Nanopore technology. This study confirms that Illumina and Nanopore sequence data provide comparable DST results and isolate cluster-identification.

## Introduction

Ten years of progress in reducing the global burden of tuberculosis (TB) have likely been lost due to the SARS-CoV-2 pandemic, with 1·4 million fewer patients diagnosed and treated in 2020 than in 2019.^1,2^ Accurate diagnosis and appropriate treatment are key to setting the global effort to end TB back on course.^3^ Understanding and interrupting transmission are equally important, as is implementing appropriate therapy for every patient. In high-income settings, whole-genome sequencing (WGS) has become an attractive solution to both these challenges, with some settings now relying predominantly on WGS for drug susceptibility testing (DST) and implementation of individualised therapeutic regimens,^4^ in addition to the well documented benefits of using these data for surveillance and outbreak control.^5^

The necessary high capital outlay for sequencing platform acquisition has limited access to TB WGS analyses in many high-burden, low-income settings. With the availability of multiple DNA sequencing platforms, simplified access to interpretation of sequencing data^5^ and curated genomic databases,^6^ more countries are now integrating DNA sequencing within TB control programs for either or both DST and epidemiological surveillance.^7^

Illumina sequencing platforms are the market leaders and are the established reference standard for TB genomic next generation sequencing (NGS). Per-base sequencing accuracy is extremely high, making this an attractive tool for both susceptibility testing and for surveillance, where just a few erroneous basecalls can be the difference between triggering public health interventions, or not. Significant validation and accreditation work has led to integration of this technology within routine clinical diagnostics in some settings (e.g. UK, the Netherlands, New York State).

Illumina technology requires large capital outlay and significant testing volume to ensure clinically appropriate turn-around times while remaining cost-efficient. Oxford Nanopore Technologies (ONT) offer a more transportable solution in the form of their handheld MinION sequencing platform (referred to as Nanopore henceforth). To date, a major obstacle for ONT’s technology has been its basecalling error rate. However, as the technology has matured and its basecalling software has improved, it is now increasingly integrated in public health laboratories.^8,9^ Although laboratory infrastructure is still required for TB culture, DNA extraction, and library preparation, the Nanopore sequencing platform’s inherent portability and relatively lower cost represent a clear opportunity for settings to benefit from WGS technology where it has hitherto been out of reach.

To date, a few studies have evaluated the accuracy of Nanopore-based genotypic DST.^9–12^ However, the impact of this sequencing technology on the clustering of isolates in the context of TB outbreak investigation remains poorly understood. We now take the opportunity to compare its performance to Illumina platforms in a head-to-head study and assess whether the accuracy of its outputs has improved sufficiently to justify its use for patient care and public health.

## Methods

### *Mycobacterium tuberculosis* clinical isolates

The *M. tuberculosis* isolates used in this study were selected in three distinct countries. In Madagascar (n=109), isolates consecutively referred to the national drug resistance surveillance program and confirmed as multi-drug resistant TB by culture were retrospectively included together with a 1:1 matched drug susceptible sample from the same sampling dates and geographical region. Ten patient samples had a second isolates taken 2 months later as per the National Tuberculosis Program “test of cure” recommendation. From South Africa (n=67), biobanked clinical isolates were selected from patients routinely diagnosed with rifampicin-resistant TB in the Western Cape Province. From England’s National Mycobacteria Reference Service in Birmingham (n=32) samples were selected from routine sequencing of mycobacterial isolates, for a total of 208 isolates. In all locations this study involved only accessing stored bacterial cultured isolates, and not directly obtaining or processing human samples. See Supplementary Section S1 for further details of isolate selection.

### Genomic sequencing, data preparation and quality control

Each isolate was sequenced on both Nanopore and Illumina platforms using extracted DNA from the same bacterial culture. See Suppl. Sections S1 for detailed DNA extraction methods, S2 for detailed sequencing methods, and S3 for human-read removal and quality control methods. After decontamination by aligning reads to a database of contaminants, data with mean read depth less than 20/30 (Illumina/Nanopore) were excluded from the study.13

### Variant calling

Illumina single nucleotide polymorphism (SNP) calls were made using the COMPASS pipeline^13^ (https://github.com/oxfordmmm/CompassCompact) used by the United Kingdom Health Security Agency (UKHSA).^7^ Nanopore SNP calls were made using BCFtools (v1.13).^14^ See Suppl. Section S4 for details of variant calling and filtering.

### Evaluation of variant call precision and recall

We evaluated the precision and recall of the SNP calls for isolates with PacBio “truth” assemblies (n=7, see Suppl. Section S5). Here, precision and recall are defined as the proportion of SNP calls that are true positives and the proportion of expected (“true”) SNP calls correctly identified, respectively. For this work, we optimise for precision following the policy of UKHSA/COMPASS.

### Assessing clusters based on SNP thresholds

We used the pairwise distance matrix to assess the impact of sequencing technologies on commonly used SNP thresholds for isolate clustering. For a given SNP threshold *t*, we constructed a clustering network (graph) by connecting isolates with a distance ≤ *t*. That is, a cluster is a subgraph within which a path exists between any two isolates, but no path exists to any isolates in another cluster. With this definition, all clusters have a minimum of two members. Isolates that do not cluster with any others are deemed singletons.

As we seek to show concordance of Nanopore data with UKHSA’s Illumina-based strategy, we investigated SNP threshold values 5 and 12.^15^ Our goal was to establish whether Nanopore data can be used to reproduce equivalent clusters to those generated with Illumina data. We therefore treat Illumina as the established standard when comparing clustering, and consider Illumina clusters as “truth”. We define three metrics (formal definitions in Suppl. Section S6 and an illustrated example in Suppl. Section S7). *Sample-averaged cluster recall* (SACR) indicates whether isolates have been missed by Nanopore clustering (false negatives) and *sample-averaged cluster precision* (SACP) reflects additional isolates being clustered by Nanopore (false positives). SACR and SACP do not account for Nanopore clusters composed solely of Illumina singletons, so we define the *excess clustering rate (*XCR) as the proportion of Illumina singletons that are clustered by Nanopore. A value of 0·1 would indicate that 10% of Illumina singletons were part of a Nanopore cluster.

### Simulation of isolate clusters with mixtures of sequencing modalities

To model the impact of using distinct sequencing platforms when supporting epidemiological investigations, we simulated mixed technology datasets by randomly choosing a technology for each isolate. We use Nanopore-to-Illumina ratios 0·01, 0·05, 0·1, 0·25, 0·5, 0·75, and 0·9. For each ratio and SNP threshold combination we performed the following 1000 times: i) randomly assign isolates to a technology in the relevant ratio, ii) calculate SACR, SACP, and XCR for the relevant SNP threshold.

### Phenotypic drug susceptibility testing

Phenotypic DST data was generated by Madagascar and South African laboratories according to local routine protocols (see Suppl. Section S8).^16^ In Madagascar, the indirect proportion method on Löwenstein-Jensen medium was performed to test the susceptibility of positive cultures against streptomycin, isoniazid, rifampicin, ethambutol, kanamycin, amikacin and capreomycin. The critical concentrations used were 4, 0.2, 40, 2, 30, 30, and 40 μg/ml, respectively. For the South African isolates, all phenotypic DST was done on Middlebrook 7H with concentrations 0.2, 2.0 and 4.0 μg/ml for isoniazid, ofloxacin and amikacin respectively. No phenotypic DST data was available for the English isolates as this is no longer routinely done by UKHSA.

### Drug resistance prediction from sequencing data

We used Mykrobe (v0.10.0) to obtain predictions of each isolate’s drug susceptibility profile for 11 drugs.^10^ See Suppl. Section S9 for detailed commands used. Mykrobe genotypes sequencing reads against a catalogue of resistance-conferring mutations. This process is independent of the variant calling steps outlined for isolate clustering. The catalogue of resistance mutations used by Mykrobe v0.10.0 consists of 476 SNPs defined at the amino acid level (which translates into 3,352 at the nucleotide level), 60 promoter SNPs, and 1,904 nucleotide-level SNPs, insertions and deletions (dominated by those in the rifampicin resistance determining region (RRDR) of the gene *rpoB*). In addition, in order to detect isoniazid and pyrazinamide resistance-causing frameshifts in the genes *katG* and *pncA*, the catalogue contains an explicit list of all possible 1 and 2bp frameshifts in those two genes, totalling 61,258.^10^ We chose not to use the recent mutation catalogue from the WHO^6^ -which came out towards the end of this study - as there is no Mykrobe version of it yet, and the purpose of this study is to determine whether Nanopore genotypes of resistance mutations are consistent with Illumina, which is independent of the catalogue.

## Results

There were 57 isolates that failed to pass quality control measures (see Genomic sequencing, data preparation and quality control), including insufficient depth for 44 isolates (37 Nanopore, 1 Illumina, and 6 both). A single lineage call could not be determined for 12 isolates. Additionally, one isolate was found to have non-matched Illumina and Nanopore data, likely due to a labelling mix-up. This left 151 isolates, sequenced on both Illumina and Nanopore platforms, that passed quality control – 91, 41, and 19 from Madagascar, South Africa, and England, respectively – with seven (Madagascar) having associated PacBio data.

For the 91 isolates from Madagascar, results for rifampicin (n=91), isoniazid (91), streptomycin (91), ethambutol (90), amikacin (51), kanamycin (51), capreomycin (51), and ofloxacin (51) were available. For the 41 South African isolates, amikacin (n=38), ofloxacin (36), isoniazid (8), capreomycin (1), ethambutol (1), kanamycin (1), and rifampicin (1) were available. Available drug susceptibility phenotypes are summarised in Figure 1.

**Figure 1:**
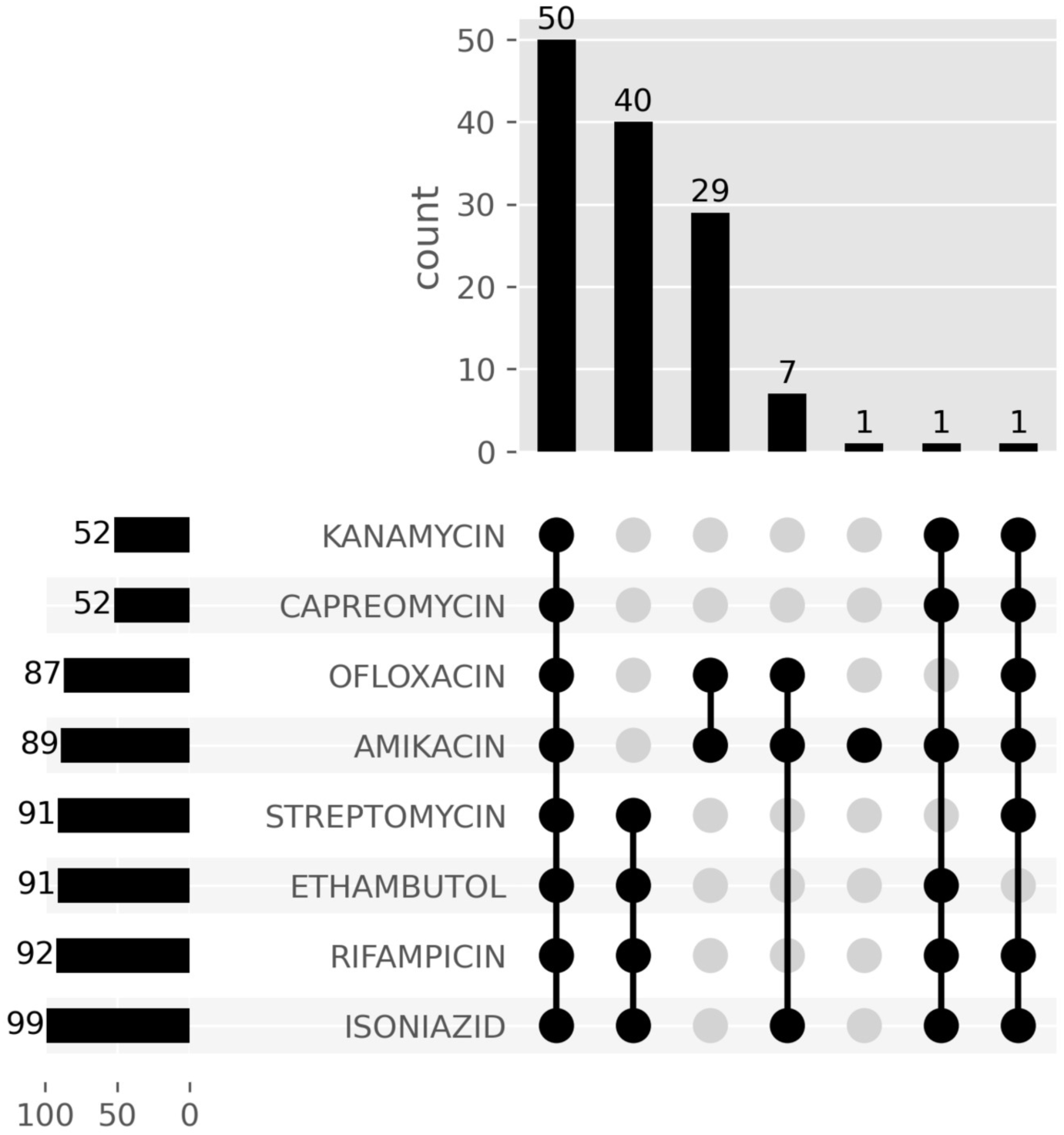
Culture-based drug susceptibility data available for isolates. Each row is a drug, and the columns represent a set of isolates that have phenotype information for those drugs with a filled cell. The top panel shows the number of isolates in the set for that combination of drugs. The bar plot in the left panel shows the number of isolates with phenotype information for each drug. Phenotypic DST was performed by clinical laboratories according to local testing algorithms which included complementary molecular testing and reflex sequential testing of second-line drugs. This explains why not all antibiotics were tested on all isolates.

### Calibrated filters achieve high accuracy

The seven isolates with PacBio truth assemblies (see Suppl. Section S5) allowed us to assess variant-calling filter thresholds and achieve different balances of precision versus recall. Since our goal was to determine whether Nanopore could be used as an alternative (or complement) to existing Illumina-based pipelines, including COMPASS used by UKHSA, we sought to match their approach, prioritising precision over recall. We show in Figure 2 the effect of applying successive filters (outlined in Variant calling). The final set of filters we used in the rest of this study are represented by the right-most boxes, leading to median SNP precision and recall of 99·5% and 90·2%, respectively, for the seven validation isolates. By comparison, Illumina data processed with COMPASS achieved median precision and recall values of 99·6% and 91·9%, respectively.

**Figure 2:**
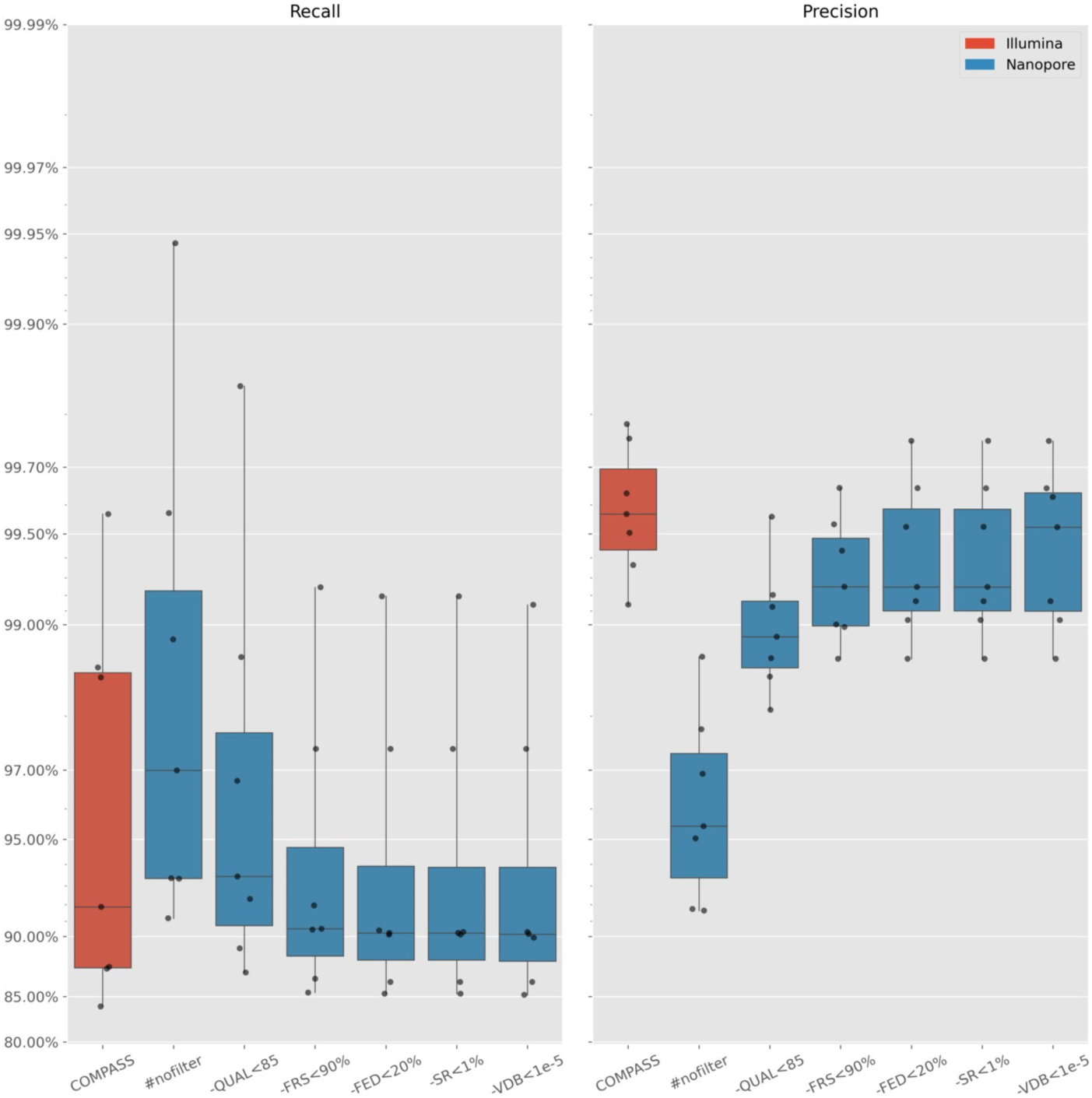
Recall (left) and precision (right) of SNPs for COMPASS/Illumina (red) and a selection of BCFtools/Nanopore filters (blue). Note the non-linear y axis scale. Each point represents a single isolate with a PacBio assembly. *#nofilter* is BCFtools with no filtering of variants. Moving right from *#nofilter*, each box accumulates a new filter plus the previous ones. Each filter describes the criterion for removing a SNP. *-QUAL<85* removes SNPs with a quality score less than 85; *-FRS<90%* removes SNPs where less than 90% of reads support the called allele; *- FED<20%* removes SNPs with read depth below 20% of the isolate’s median depth; *-SR<1%* removes SNPs with less than 1% of read depth on either strand; *-VDB<1e-5* removes SNPs with a variant distance bias less than 0·00001.

Applying these filters to all 151 study isolates, we show in Suppl. Figure S2 the SNP distances between all pairs of isolates as measured by COMPASS on the Illumina data, and our BCFtools pipeline on the Nanopore data. The distances are highly correlated, but as expected, the lower recall of the Nanopore/BCFtools pipeline results in systematically lower estimated SNP differences (many points lie below *y* = *x*). Figure 3 additionally shows the distance correlation for those isolates within 20 Illumina SNPs of each other - i.e., the isolates most relevant to transmission investigations. Encouragingly, at a distance threshold of 12, only two pairs of isolates (red points) would fail to be linked by Nanopore, although we will later show that this does not cause these isolates to be missed from their wider clustering.

**Figure 3:**
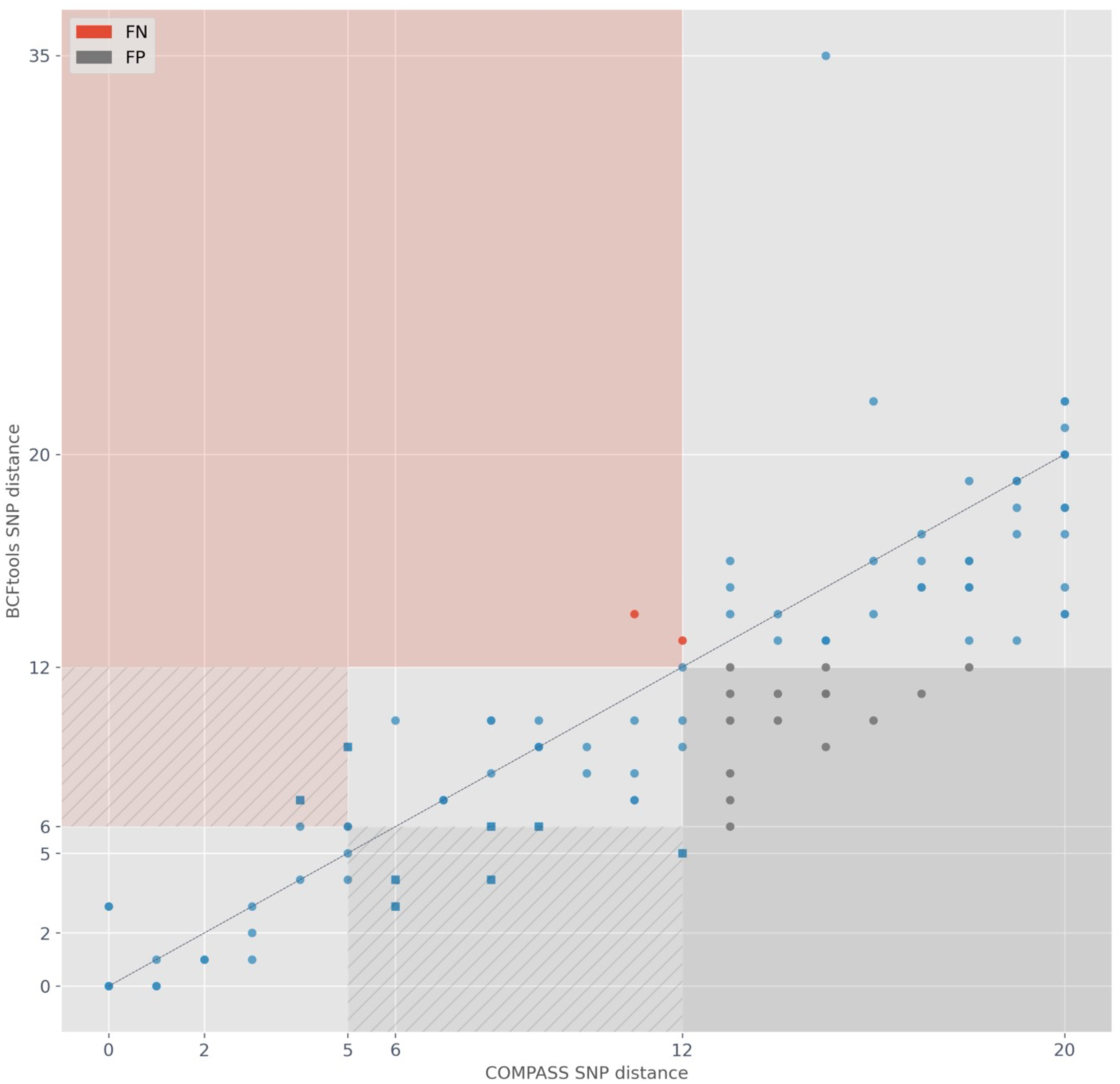
Pairwise SNP distance relationship between Illumina (COMPASS; x-axis) and Nanopore (BCFtools; y-axis) data. Each point represents the SNP distance between two isolates. The black, dashed line shows the identity line (i.e., y=x). The isolate pairs shown are all pairs where the COMPASS distance is ≤ 20. The red area and points indicate pairs with a Nanopore distance *>* 12 but an Illumina distance ≤ 12. These pairs are deemed false negative (FN) connections. The red area with stripes indicates pairs that are FN connections at an Illumina threshold of 5 (Nanopore threshold 6), but not when the threshold is expanded to 12. These pairs are shown as square points. The grey area and points are the inverse - i.e., false positive (FP) connections. Thus the grey striped area shows pairs of samples which are FP connections at an Illumina threshold of 5 (Nanopore threshold 6), but not when the threshold is expanded to 12.

### Nanopore-based transmission clusters recapitulate baseline Illumina clusters

We compare the clusters obtained from COMPASS/Illumina and BCFtools/Nanopore SNP calls using single-linkage clustering with the standard 5- and 12-SNP thresholds previously reported to correspond to highly and moderately probable transmission events.^15,17,18^ We developed three metrics (SACR, SAC, XCR) to provide a quantitative assessment of how Nanopore clusters differed from baseline Illumina ones (see Assessing clusters based on SNP thresholds).

As we see in Figure 3, Illumina and Nanopore SNP distances do not lie exactly on *y* = *x*, and so we need to use slightly different Nanopore SNP-thresholds to match Illumina results. For Illumina thresholds 5 and 12, we selected the respective Nanopore SNP threshold that gave the best balance of SACR, SACP, and XCR values (Suppl. Section S11). As we sought to minimise the number of isolates missed from their true cluster, a higher SACR is favoured. We selected Nanopore SNP thresholds of 6 and 12 (see Suppl. Figure S3). In Figure 4 we show Illumina and Nanopore clusters at these thresholds. Nodes represent isolates, coloured according to their Illumina clusters. We find isolates clustered together by Illumina remain clustered with Nanopore (nodes of the same colour are connected). At threshold 5, the Illumina clusters are recapitulated, but two Illumina-singleton isolates are adjoined to cluster 2 (dark orange), and seven Illumina-singleton isolates are combined into two new clusters of five and two isolates. At threshold 12, clusters 1 and 2, and clusters 7 and 8 are merged by Nanopore. In both cases, the Illumina maximal SNP distance between isolates included in these clusters is 13 - one above the established threshold. There are also five Illumina singletons which are adjoined to pre-existing clusters by Nanopore. Figure 4 also shows the metrics SACP, SACR, and XCR for each threshold, providing some intuition for how the values correspond to changes in clustering. Note, for example, that SACP for threshold 12 is 0·817. For both thresholds, the SACR value is 1·0, meaning Nanopore does not miss any isolates from their correct cluster. All clusters exclusively regroup isolates from a same single country. Additionally, isolates from the same patient (n=8) were also clustered together by both technologies.

**Figure 4:**
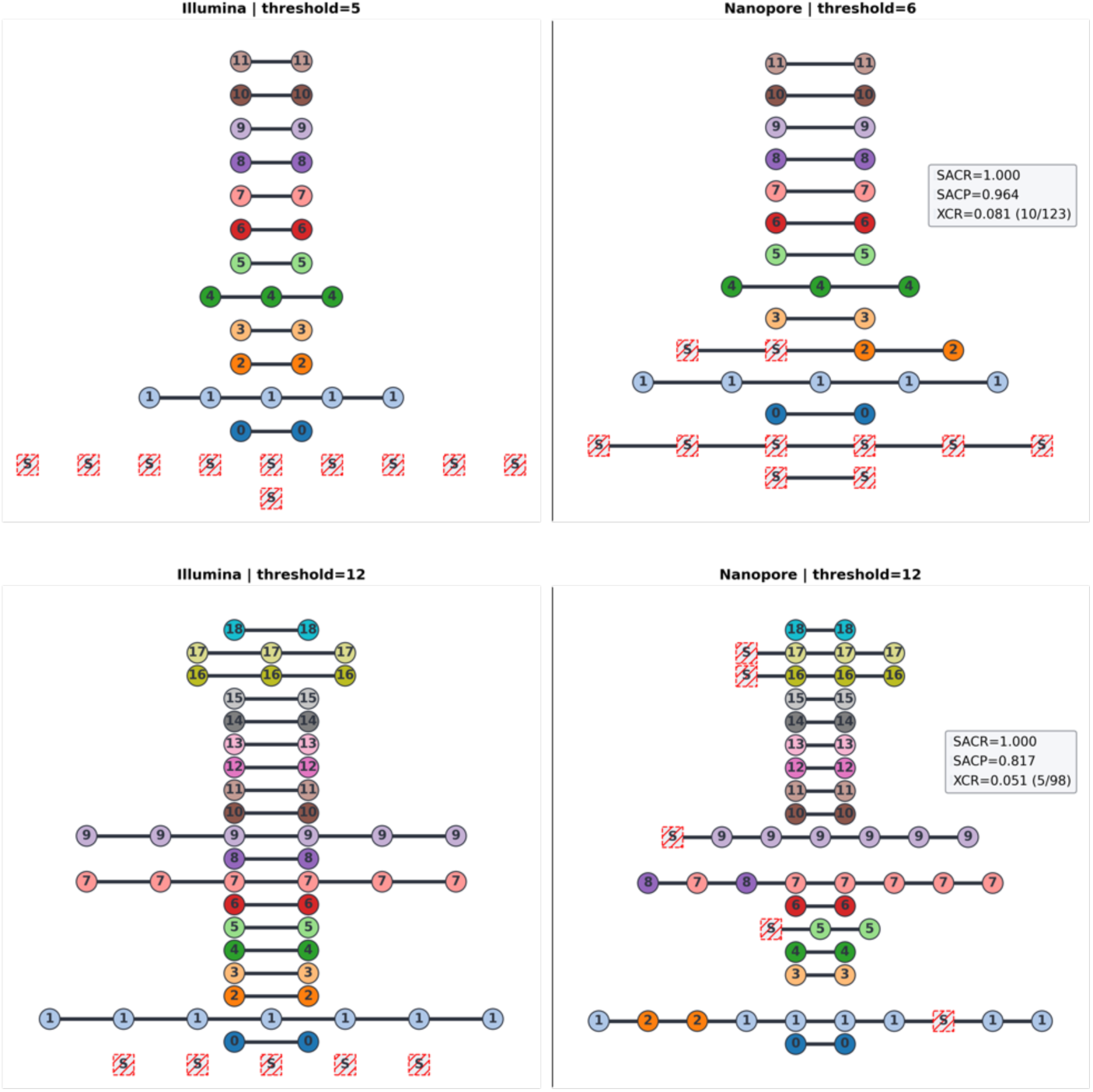
Agreement of Illumina and Nanopore transmission clustering for two thresholds of interest: 5-Illumina/6-Nanopore (upper panels) and 12-both technologies (lower panels). The expected (Illumina/COMPASS) clusters are shown in the left panels, with the Nanopore/BCFtools clustering shown on the right. The title of each panel indicates the SNP threshold used for clustering. Nodes are coloured and numbered according to their Illumina cluster membership. Isolates clustered by Nanopore and not clustered (singletons) by Illumina are represented as white boxes with red stripes and are named “S”. Clusters are horizontally aligned and connected with black lines; however, the order of nodes and the length of edges has no significance. In addition, each Nanopore panel has a legend showing the SACR, SACP, and XCR value with respect to the Illumina clustering. SACR=sample-averaged cluster recall; SACP=sample-averaged cluster precision; XCR=excess clustering rate (with the raw numbers in parentheses).

### Clustering with mixed technology data performs consistently

As an initial check, the “self-distance” was calculated - i.e., the SNP distance between the Nanopore- and Illumina-derived consensus genomic sequence for each isolate. The histogram of these values is shown in Suppl. Figure S4, confirming these distances were close to zero (mean=1·2; median=0).

Since our dataset consists of 151 isolates with both Illumina and Nanopore data, we are able to simulate a wide range of mixed technology datasets by randomly assigning either the Nanopore or Illumina data to each isolate (see Simulation of isolate clusters with mixtures of sequencing modalities). We generated 1000 simulated datasets for each value in a range of Nanopore:Illumina ratios (0·01, 0·05, 0·1, 0·25, 0·5, 0·75, and 0·9), and measured the impact on SACR, SACP, and XCR. As shown in Figure 5, as we increase the proportion of Nanopore data, the recall (SACR) behaves consistently, with median fixed at 1·0, and the precision (SACP) degrades smoothly from 1·0 (meaning near-pure Illumina data perfectly recapitulates pure Illumina clusters) to the value in the pure Nanopore dataset (0·964/0·817 for thresholds 5/12). XCR behaves in the same way as SACP; gradually decreasing to the pure Nanopore level.

**Figure 5:**
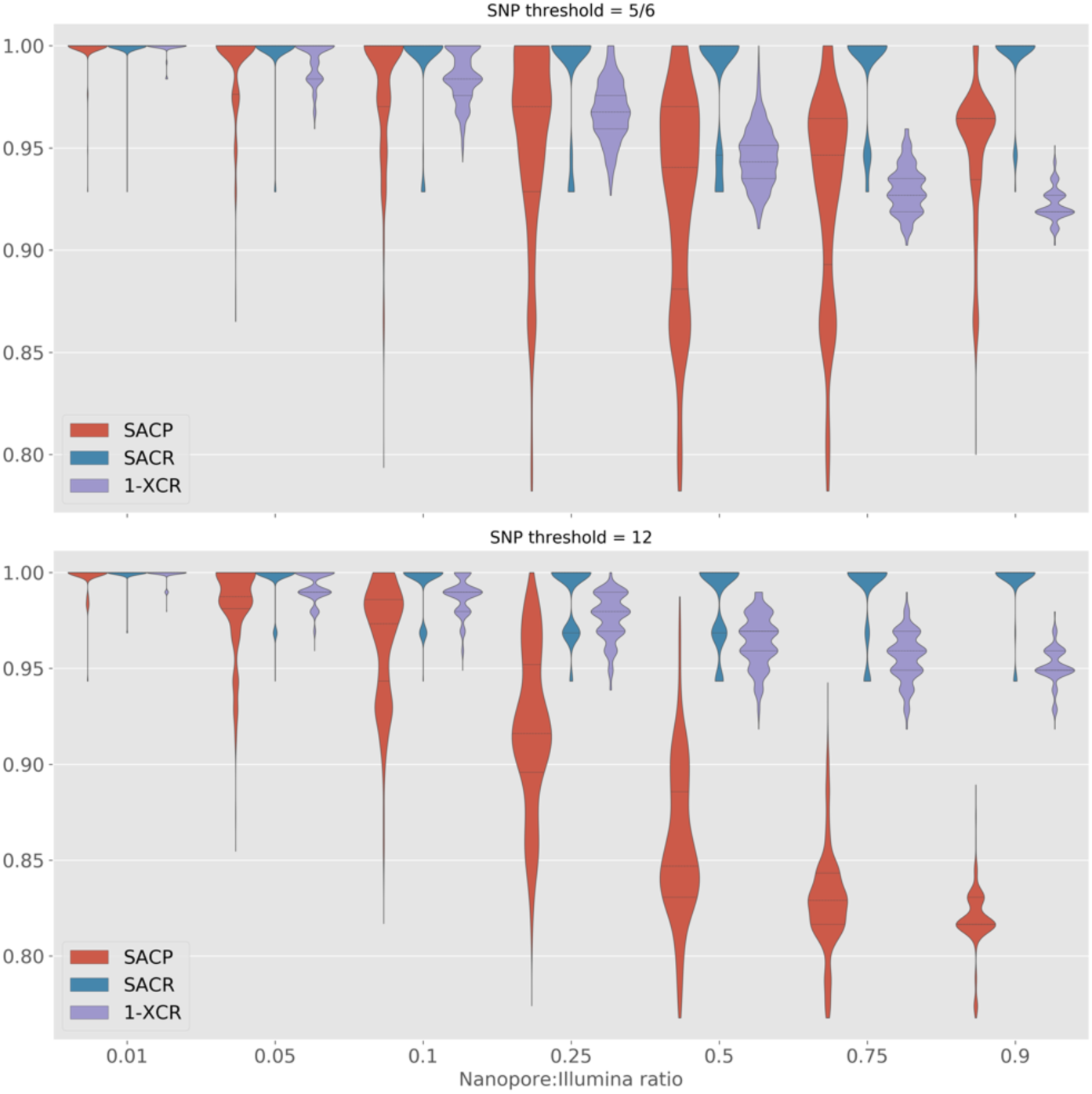
Simulating various ratios (x-axis) of Nanopore/Illumina isolate mixtures. The different thresholds (subplots) indicate the cut-off for defining isolates as part of a cluster. The y-axis depicts the sample-averaged cluster precision and recall (SACP/SACR) and excess clustering rate (XCR) distributions over all simulation runs. For each ratio/threshold combination we run 1000 simulations where the Nanopore and Illumina data is randomly split into the relevant ratio and clusters are defined based on the relevant threshold. The titles for each subplot indicate the SNP threshold used when comparing Illumina, Nanopore, or mixed-technology isolate pairs (5/6 is 5 for Illumina and 6 for Nanopore and mixed).

To give some intuition on these metrics, consider a simulated sample of 100 isolates including three clusters of size 2, 2, and 9, with 87 singletons. Half are sequenced on Nanopore and the other half on Illumina. The expected SACR/SACP/XCR from the simulations in this section are 1·0/0·847/0·031 for a SNP threshold of 12. The recall (SACR) suggests we would expect all isolates in our hypothetical dataset to be clustered with their expected neighbours. The precision (SACP) of 0·847 in this example would be equivalent to the 2 two-member clusters being joined into a single cluster, while an XCR value of 0·031 could be caused by 3 singleton isolates forming a new cluster.

### Nanopore-based genotyping of resistance mutations is highly concordant with Illumina

We compared the Nanopore and Illumina genotype calls of Mykrobe at the 66,537 nucleotide-level resistance conferring mutations for our 151 isolates (see Drug resistance prediction from sequencing data). In total, we found four genotype discordances. Three of these discrepant mutations were *katG* 1bp deletions at consecutive positions within a homopolymer in *katG*, all in the same isolate, effectively describing one deletion event - thus only affecting a single phenotype call. The other discrepancy was a *katG* 1bp deletion in a separate isolate. (There were also two further mutations (each in one isolate) which we did not classify as discrepant, where a resistance mutation was detected with both Nanopore and Illumina, but filtered in the Illumina calls due to low coverage (*rrs* a1401g and *rrs* a514c)). For further details see Suppl. Section S9.1. A summary of concordance of predictions is shown in Table 1. These results lead to the key observation for evaluating the utility of Nanopore data as a replacement or complement for Illumina for obtaining genotypic DST: if genotyping at resistance mutations is highly concordant (here 100% for SNPs and >99.99% for indels), then this concordance should be retained as catalogues of resistance mutations are improved.

**Table 1:**
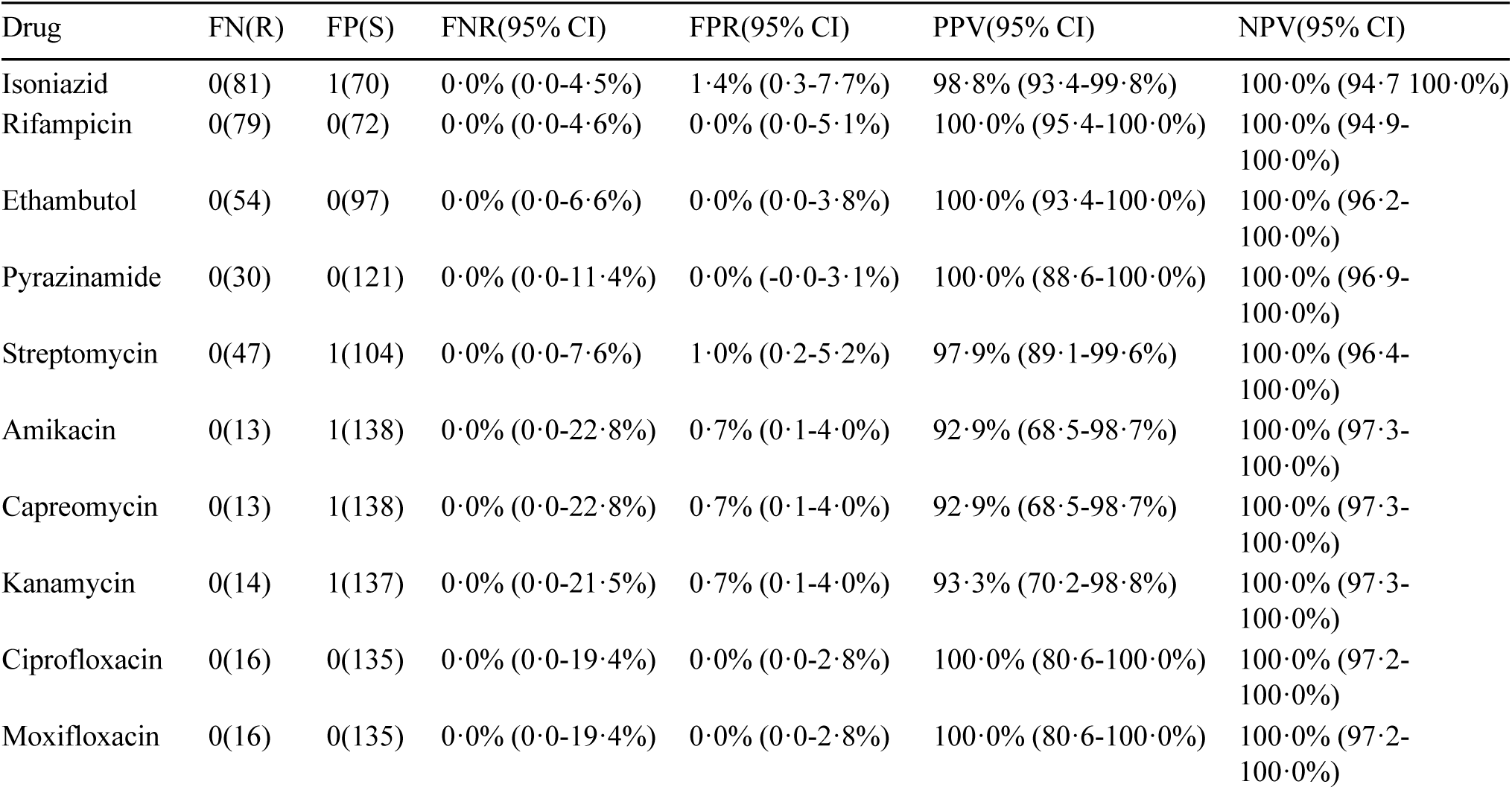
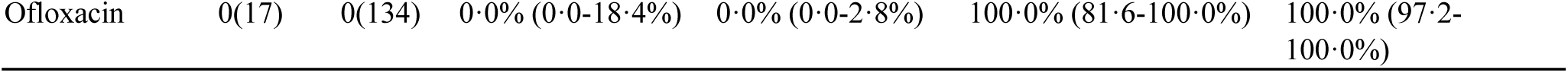
Comparison of Mykrobe-derived Nanopore drug resistance predictions with Illumina predictions. For this comparison, we consider the Mykrobe resistance prediction from Illumina as the reference standard. FN=false negative, meaning Nanopore does not detect resistance where Illumina does; R=number of resistant isolates; FP=false positive, meaning Nanopore detects resistance where Illumina finds susceptible; S=number of (Illumina) susceptible isolates; FNR=false negative rate; FPR=false positive rate; PPV=positive predictive value; NPV=negative predictive value; CI=Wilson score confidence interval.

| Drug | FN(R) | FP(S) | FNR(95% CI) | FPR(95% CI) | PPV(95% CI) | NPV(95% CI) |
| --- | --- | --- | --- | --- | --- | --- |
| Isoniazid | 0(81) | 1(70) | 0.0% (0.0-4.5%) | 1.4% (0.3-7.7%) | 98.8% (93.4-99.8%) | 100.0% (94.7-100.0%) |
| Rifampicin | 0(79) | 0(72) | 0.0% (0.0-4.6%) | 0.0% (0.0-5.1%) | 100.0% (95.4-100.0%) | 100.0% (94.9-100.0%) |
| Ethambutol | 0(54) | 0(97) | 0.0% (0.0-6.6%) | 0.0% (0.0-3.8%) | 100.0% (93.4-100.0%) | 100.0% (96.2-100.0%) |
| Pyrazinamide | 0(30) | 0(121) | 0.0% (0.0-11.4%) | 0.0% (-0.0-3.1%) | 100.0% (88.6-100.0%) | 100.0% (96.9-100.0%) |
| Streptomycin | 0(47) | 1(104) | 0.0% (0.0-7.6%) | 1.0% (0.2-5.2%) | 97.9% (89.1-99.6%) | 100.0% (96.4-100.0%) |
| Amikacin | 0(13) | 1(138) | 0.0% (0.0-22.8%) | 0.7% (0.1-4.0%) | 92.9% (68.5-98.7%) | 100.0% (97.3-100.0%) |
| Capreomycin | 0(13) | 1(138) | 0.0% (0.0-22.8%) | 0.7% (0.1-4.0%) | 92.9% (68.5-98.7%) | 100.0% (97.3-100.0%) |
| Kanamycin | 0(14) | 1(137) | 0.0% (0.0-21.5%) | 0.7% (0.1-4.0%) | 93.3% (70.2-98.8%) | 100.0% (97.3-100.0%) |
| Ciprofloxacin | 0(16) | 0(135) | 0.0% (0.0-19.4%) | 0.0% (0.0-2.8%) | 100.0% (80.6-100.0%) | 100.0% (97.2-100.0%) |
| Moxifloxacin | 0(16) | 0(135) | 0.0% (0.0-19.4%) | 0.0% (0.0-2.8%) | 100.0% (80.6-100.0%) | 100.0% (97.2-100.0%) |
| Ofloxacin | 0(17) | 0(134) | 0·0% (0·0-18·4%) | 0·0% (0·0-2·8%) | 100·0% (81·6-100·0%) | 100·0% (97·2-100·0%) |

### Concordance with phenotypic DST (limited by catalogue, not technology)

Finally, for completeness, we show the agreement of WGS predictions with culture-based phenotype. Figure 6 and Suppl. Table S2 show the results for all isolates and drugs with available DST phenotypes. As expected, we see the Nanopore and Illumina results are nearly identical. Nanopore produced 2 fewer missed resistance (FN) calls than Illumina (amikacin and streptomycin). However, Nanopore data lead to one extra false resistance (FP) call compared to Illumina (isoniazid). In addition, we found there was no relationship between low (Nanopore) read depth and reduced prediction performance (see Suppl. Section S14) - indicating we saw no improvement in prediction accuracy with high sequencing depth (>30x).

**Figure 6:**
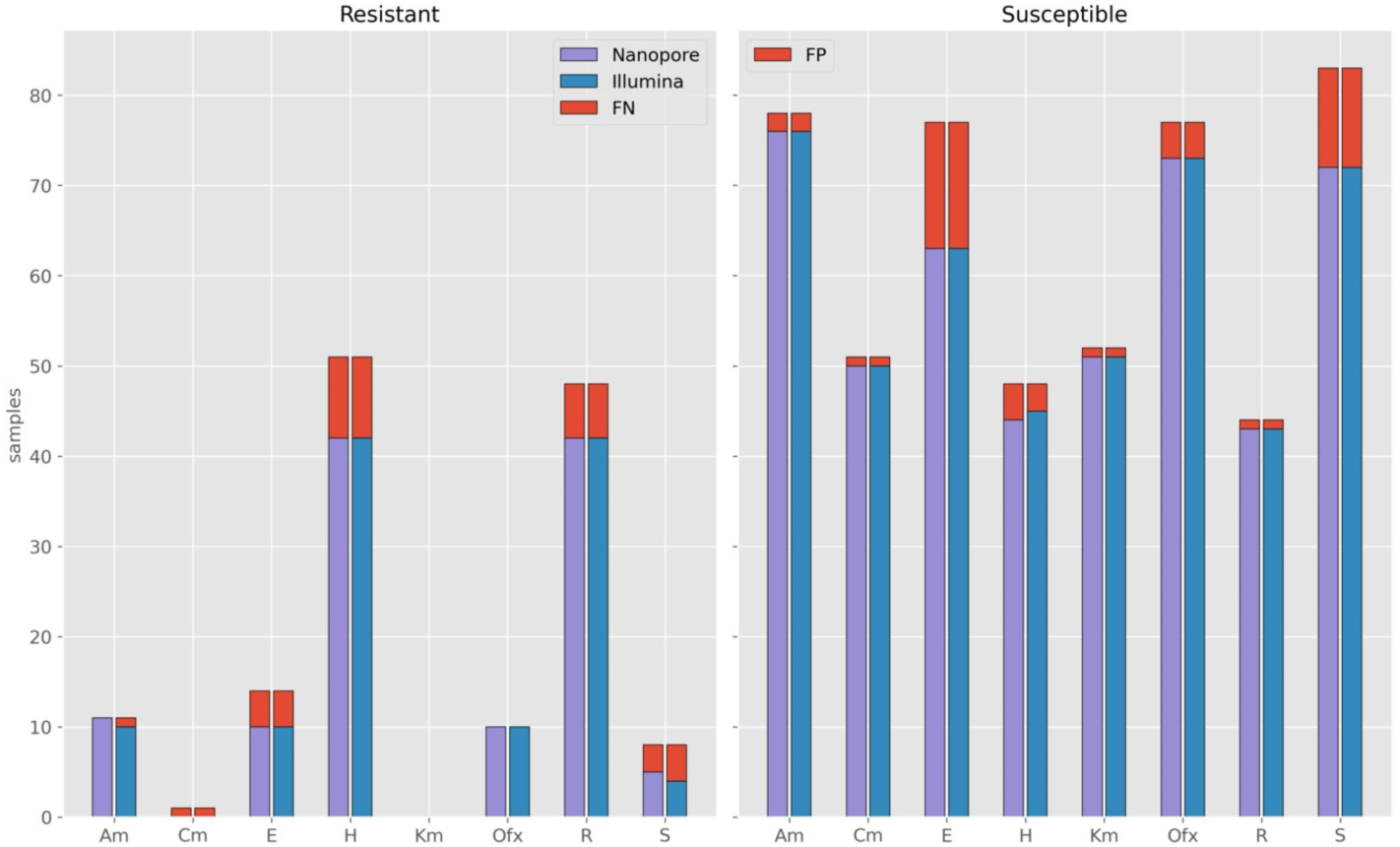
Number of resistant (left) and susceptible (right) phenotypes correctly predicted by Mykrobe from Illumina (blue) and Nanopore (purple) WGS data. The red bars indicate missed (FN) or incorrect (FP) predictions. The x-axis shows the drugs with available phenotype data. Am=amikacin; Cm=capreomycin; E=ethambutol; H=isoniazid; Km=kanamycin; Ofx=ofloxacin; R=rifampicin; S=streptomycin.

## Discussion

The need for precision diagnostics supporting TB DST and transmission interruption is imperative. There is an increasing range of settings in which *M. tuberculosis* genomic sequencing is deployed. The added value of this technology depends on local TB incidence, including that of drug-resistant infections and local disease transmission patterns. Although the resistance and transmission use cases may motivate the adoption of TB WGS, operational characteristics, costs, and analytical performance need consideration when committing to a sequencing platform. This paper compares the established Illumina and emerging Nanopore technologies in their ability to address the most pragmatic questions TB clinicians and control officers use bacterial genomic sequencing for: predicting drug resistance and identifying putative transmission clusters using SNPs.

These tasks rest on two pillars. First, how well understood are the genetic determinants of resistance to the various antitubercular drugs and how can they be used to predict resistance from genotype. Second, how well the sequence data from an isolate can be “assayed” to either detect all SNPs (used for clustering) or evaluate a list of known polymorphic positions (used for genotypic DST). The first question is technology-independent and has been the subject of many studies over recent years. Multiple studies have compiled catalogues of resistance mutations,^4,10,11,19^ and recently the WHO has published a knowledgebase of high confidence mutations intended to provide a solid foundation for future catalogues.^6^ Given perfect sequencing, the catalogue determines how well DST can be predicted, which is inexorably improving as the global community collects progressively more data.^20–23^ In this study we take this as given, and ask whether Nanopore sequence data can provide as accurate genotyping of the resistance catalogue as Illumina data. If so, as catalogues improve, both Illumina and Nanopore data will provide concordant and progressively better results.

Our analysis shows that it is now possible to obtain high-precision SNP calls in *M. tuberculosis* with current Nanopore data, with only a limited decrease in recall - we obtain median precision/recall of 99·5/90·2% with Nanopore data, compared with 99·6/91·9% for Illumina. These translate into 5- and 12-SNP clusters congruent with those produced by Illumina data. One can be highly confident that isolates will not be omitted from “true” transmission clusters - i.e., those that Illumina data would deem clustered. We find that where Nanopore over-clusters compared with Illumina, the additional isolates are closely related and just beyond the Illumina threshold. In terms of genotyping resistance-causing SNPs and indels, the two technologies give almost identical results using Mykrobe - four discordances amongst 151 isolates x 66,537 nucleotide-level resistance-conferring mutations in the catalogue gives a concordance >99.99%. We conclude that (given >30x depth) Nanopore data can generate functionally equivalent data to Illumina for our specific pragmatic goals (resistance prediction and cluster detection).

These results extend and improve on recent work evaluating Nanopore sequencing for *M. tuberculosis* clinical applications.^9,24^ In Smith *et al*., Illumina/Nanopore concordance at resistance SNPs was measured as 98·1% across 431 isolates.^9^ However SNP accuracy was not measured (as that needs a gold standard), and cluster concordance was not evaluated at all. This study fills those gaps: producing a high-quality truth set based on PacBio sequencing enabling us to measure ground truth as well as concordance; performing a thorough evaluation of clustering using several novel metrics; an explicit assessment of mixed technology datasets on isolate clustering; and providing reproducible software workflows on which others can build.

Our study, by design, did not include detailed conventional epidemiology data to compare with the molecular clusters. We acknowledge that pure SNP-based threshold clustering has shortcomings. Stimson *et al*. recently published a notable study showing that combining a SNP threshold approach with epidemiological data can lead to superior transmission cluster reconstruction than SNP threshold alone.^25^ The work we present here lays a foundation for investigating how Nanopore data performs with these more nuanced approaches.

There has been a remarkable continual evolution and improvement of Nanopore data quality over the last five years, which appears to continue unabated. We found our results evolved over the course of the study as basecalling software and BCFtools were updated (data not shown). This required careful recalibration of variant filters. Newer R10 flow cells will likely also require recalibration of filters. As such, we strongly encourage validation of new software versions and flow cells – particularly basecalling and variant calling - and note that the data we present provides a valuable test set for this quality control.

## Contributors

All authors meet criteria for authorship as per the Lancet Infectious Diseases policy and ICJME recommendations. Conceptualization (MBH, SGL, ZI), *M. tuberculosis* culture and sequencing (MSR, MG, SoG, RW, SeG, DC, SGL), data analysis plan (MBH, AK, AD, NR, HC, SGL, ZI), bioinformatics, formal analyses, visualisations, and software design (MBH), writing of the original draft (MBH, SGL, ZI), funding acquisition (TW, SGL). All authors have read, reviewed, discussed, and approved the final manuscript and their respective representation in the authorship.

## Data Availability

All data produced in the present work are contained in the manuscript

## Data sharing

The code used to perform all analyses and visualisations in this study is available at https://github.com/mbhall88/head_to_head_pipeline.

All sequencing reads generated for this study have been deposited in the European Nucleotide Archive (ENA) under the Project accession PRJEB49093. A table listing accessions for each Sample and Run can be found at https://doi.org/10.6084/m9.figshare.19304648.

## Conflicts of interest

ZI, SGL and NR had travel and accommodation costs reimbursed when speaking at an Oxford Nanopore Technology (ONT) conference in 2017. SGL and NR previously received consumables from ONT when establishing Nanopore sequencing capacity in Madagascar. ONT matched the contributions from the Longitude Prize Discovery Award to ZI, TW in 2017 to provide consumables for sequencing in Vietnam and India. ONT did not provide funding (direct or in kind) for this project, and had no input or knowledge of the design, data analysis or paper writing. Funders had no input into the design, data analysis or paper writing of this project.

## Ethics

No human data was involved in this study, and therefore

## Acknowledgements

We thank the laboratory personnel of the Mycobacteria Unit of the Institute Pasteur Madagascar and the National TB program of Madagascar. We also thank the Translational Genomics Research Institute (David Engelthaler), Centre for Disease Control and Prevention (James Posey) for the contribution of a small subset of the Illumina WGS of clinical isolates from South Africa (n=7 each). SGL is supported by the *Fond de Recherche Santé Québec*. TMW is a Wellcome Trust Clinical Career Development Fellow (214560/Z/18/Z). AK was supported by a Carnegie Corporation Developing Emerging Academic Leaders Early Career Fellowship. AK, MG and RW acknowledge support from the Tuberculosis Omics Research Consortium, headed by Prof Annelies Van Rie, funded by the Research Foundation Flanders (FWO), under grant No. G0F8316N (FWO Odysseus).

This research was funded in whole, or in part, by the Wellcome Trust [214560/Z/18/Z; ISSF 204826/Z/16/Z]. For the purpose of open access, the author has applied a CC BY public copyright licence to any Author Accepted Manuscript version arising from this submission.

## Notes

### Funding Statement

SGL is supported by the Fond de Recherche Sante Quebec. TMW is a Wellcome Trust Clinical Career Development Fellow (214560/Z/18/Z). AK was supported by a Carnegie Corporation Developing Emerging Academic Leaders Early Career Fellowship. AK, MG and RW acknowledge support from the Tuberculosis Omics Research Consortium, headed by Prof Annelies Van Rie, funded by the Research Foundation Flanders (FWO), under grant No. G0F8316N (FWO Odysseus). This study was funded inpart by by the Wellcome Trust [214560/Z/18/Z; ISSF 204826/Z/16/Z].

