## Supplementary Material for "Nanopore sequencing for *Mycobacterium tuberculosis* drug susceptibility testing and outbreak investigation"

### Supplementary information

#### Contents


**S1 Isolate selection and DNA extraction** 4

**S2 Extended sequencing methods** 5

**S3 Sequencing data preparation and quality control** 7

**S4 Variant calling** 7

**S5 Consensus genomic sequence assembly** 8

**S6 Extended clustering metrics definitions** 9

**S7 Illustrated example of clustering metrics** 11

**S8 Drug susceptibility testing methods** 12

**S9 Drug resistance predictions with Mykrobe** 13

**S10 Full dataset distance plot** 14

**S11 Selecting Nanopore SNP distance thresholds** 16

**S12 Mixed modality self-distance** 18

**S13 Mixed sequencing modality distance plot** 20

**S14 Effect of Nanopore read depth on drug resistance predictions** 22

**S15 Genotypic drug resistance prediction concordance with phenotype** 24

**References** 26

### List of Figures


S1 Illustrative examples of transmission clustering. **a)** represents truth clusters, while **b)** is clustering from some "test" method we would like to compare to **a**. The nodes represent samples with the numbers on the edges connecting them indicating the distance between those two samples. The red nodes indicate samples with a clustering disparity between the two clusterings. Note, we do not show singletons (disconnected nodes) - e.g., $J$ is missing from **(b)**. . . . . . . . . . . . . . . . . . . . . . . . . . . . . . . . . . . 12

S2 Pairwise SNP distance relationship between Illumina (COMPASS; x-axis) and Nanopore (`bcftools`; y-axis). Each point represents the SNP distance between two isolates. The black, dashed line shows the identity line (i.e. $y = x$). . . . . . . . . . . . . . . . . . 15

S3 Illumina and Nanopore (`bcftools`) transmission cluster similarity for various SNP distance threshold. Each subplot compares the Nanopore clustering for the threshold on the x-axis to the Illumina clustering based on the distance (threshold) in the subplot title. SACR (red), SACP (blue), and 1−XCR are represented by the solid, dashed, and dotted lines, respectively. SACR=sample-averaged cluster recall; SACP=sample-averaged cluster precision; XCR=excess clustering rate. . . . . . . . . . . . . . . . . . . . . . 17

S4 Mixed modality "self-distance". This plot shows the SNP distance (x-axis) between each isolate's COMPASS (Illumina) and `bcftools` (Nanopore) VCF calls. Note, the y-axis is log-scaled. 19

S5 Pairwise SNP distance relationship between Illumina (COMPASS; x-axis) and mixed COMPASS-`bcftools` calls (y-axis). Each point represents the SNP distance between two isolates. The black, dashed line shows the identity line (i.e., y=x). The zoomed inset shows all pairs where the COMPASS distance is $\leq 20$. The red area and points indicate pairs with a mixed distance $\geq 12$, but an Illumina distance $\leq 12$. These pairs are deemed false negative (FN) connections. The red area with stripes indicates pairs that are FN connections at an Illumina threshold of 5 (mixed threshold 6), but not when the threshold is expanded to 12. These pairs are shown as square points. The grey area and points is the inverse - i.e., false positive (FP) connections. Note, the y-axis in the inset window is log-scaled. . . . . . . . . . . . . . . . . . . . 21

S6 Effect of Nanopore read depth on Mykrobe DST prediction. Each point indicates the proportion (left y-axis) of classifications of that type at the read depth on the x-axis. The blue bars indicate the number of samples (right y-axis) contained in each bin. **(a)** read depth, $d$, is binned such that 40 is all samples where $40 \leq d \leq 50$. **(b)** read depth bin 40 is all samples where $d \geq 40$. FP=false positive (red); TP=true positive (blue); FN=false negative (purple); TN=true negative (grey). . . . . . . . . . . . . . 23

#### List of Tables

S1 Cluster recall and precision results for each sample in Figure S1. 12
S2 Comparison of WGS-based drug resistance predictions with culture-based phenotypes. For this comparison, we use phenotypic DST as the reference standard and evaluate Mykrobe Illumina and Nanopore resistance predictions accordingly. FN=false negative, meaning Nanopore does not detect resistance where Illumina does; R=number of resistant isolates; FP=false positive, meaning Nanopore detects resistance where Illumina finds susceptible; S=number of (Illumina) susceptible isolates; FNR=false negative rate; FPR=false positive rate; PPV=positive predictive value; NPV=negative predictive value; CI=Wilson score confidence interval. . . . . . . . . . . . . . . . . . . . . . . . . . . . . . . . . . 25
S3 Discordances between Mykrobe genotype calls, and Mykrobe predicted phenotype when comparing Illumina and Nanopore. In the genotype column the Illumina and Nanopore genotype calls are shown separated by a forward-slash Illumina/Nanopore. F = call was filtered out; 0 = reference allele; 1 = alternate allele. The "Genotype discordant" column shows whether this difference was considered a discordance in our calculation. The "Phenotypes (predicted)" column shows the Mykrobe predictions for Illumina/Nanopore: S = susceptible; R = resistant. The "Phenotype discordant" column shows whether the predicted phenotypes in the previous column are the same. *The isolate had other mutations in *katG* that cause isoniazid resistance. †We skip mutations where either technology's call is filtered (see Section S9.1). 25

### S1 Isolate selection and DNA extraction

##### Madagascar

The Institut Pasteur de Madagascar hosts the Madagascar Programme National de Lutte Contre la Tuberculose (National TB Program), which provides reference DST for TB patients suspected of facing infection relapse or treatment failure. Culture-confirmed multi-drug resistant (MDR) isolates were included. All MDR isolates were matched with culture-confirmed drug-susceptible isolates referred during the same period and from the same region of the country. All available isolates were included for patients undergoing TB treatment and for which a follow-up culture was positive.

DNA from *M. tuberculosis* culture was extracted using the cetyltrimethylammonium bromide (CTAB) method previously described by Van Embden with minor modification[1]. Briefly, samples were inactivated by heating at 80°C for 30 minutes. Next, 10mg/ml of lysozyme was added to the suspension and incubated for at least one hour at 37°C. After adding proteinase K 10mg/ml and sodium dodecyl sulphate (SDS) 10%, the suspension was incubated for 10 minutes at 65°C. A mixture of CTAB and 5M NaCl preheated to 65°C was then added. The suspension was mixed until a milky mixture was obtained and incubated for 10 minutes at 65°C. Next, chloroform isoamyl alcohol (24:1) mixture was added, followed by centrifugation for 5 minutes at 10,000rpm and 4°C. The upper phase was recovered, and DNA was precipitated by adding isopropanol to the solution. The mixture was frozen for at least one hour then centrifuged at 10000rpm and 4°C, for 5 minutes. The supernatant was discarded, and the pellet was washed with 70% ethanol and centrifuged at 10,000rpm and 4°C, for 5 minutes. The DNA pellet was dried with speed-vac for 2 minutes and resuspended in 1X TE. DNA was quantified using the Qubit dsDNA HS Assay Kit (Thermo Fisher Scientific, USA).

##### South Africa

Clinical *M. tuberculosis* isolates routinely collected in the Western Cape Province of South Africa, processed by the National Health Laboratory Service (NHLS) and diagnosed as rifampicin-resistant tuberculosis, are biobanked at the South African Medical Research Council Centre for Tuberculosis Research (SAMRC-CTR) housed at the Division of Molecular Biology and Human Genetics at Stellenbosch University, South Africa. A convenience data set of 82 clinical *M. tuberculosis* isolates for which Illumina WGS data was available were selected for Nanopore sequencing.

Clinical *M. tuberculosis* isolates were cultured on supplemented 7H10 solid media under BSL3 conditions. Phenol-chloroform DNA extraction was performed on heat-inactivated cultured isolates as previously described[2].

**England**

Total DNA was extracted from heat-inactivated, saline-washed Mycobacteria growth indicator tube (MGIT) cultures using two rounds of mechanical cell disruption, followed by DNA purification with 1x volume Agencourt AMPure XP beads, as described previously[3].

### S2 Extended sequencing methods

#### S2.1 Overview

Illumina sequencing was performed as per the manufacturer's instruction on either the MiSeq, HiSeq 2500, or NextSeq500 platforms. Nanopore sequencing was performed using the Ligation Sequencing Kit 1D (SQK-LSK108 or SQK-LSK109) and the Native Barcoding Kit 1D (EXP-NBD103 or EXP-NBD104) according to the manufacturer's instructions on either the MinION or GridION platform with R9.4.1 flow cells. In addition, 35 Malagasy isolates including drug-resistant strains and were sequenced on the PacBio CCS platform.

#### S2.2 Illumina

**Madagascar**

Illumina sequencing was carried out on the HiSeq 2500 platform at the Wellcome Trust Centre for Human Genetics, Oxford, and paired-end libraries were prepared according to the manufacturer's instruction.

**England**

Illumina sequencing was performed on a MiSeq instrument at Public Health England (Birmingham) by Grace Smith, Esther Robinson and their team. Sample preparation and sequencing methodology were as described previously[3].

**South Africa**

Paired-end genomic libraries were prepared using the Illumina Nextera XT library or NEBNext Ultra TM II FS DNA Library Preparation Kits (Illumina Inc, San Diego, CA, USA) according to the manufacturers' instructions. Pooled samples were sequenced on an Illumina HiSeq2500 or NextSeq500 instrument.

#### S2.3 Nanopore

**Madagascar**

Nanopore library preparation was carried out using the Oxford Nanopore Technology (ONT) Ligation Sequencing Kit 1D (SQK-LSK108) and the Native Barcoding Kit 1D (EXP-NBD103) according to the ONT standard protocols. One microgram of DNA was used as input for each library. Multiplexed sequencing was performed by pooling 6-8 barcoded DNA samples. Prepared libraries were loaded onto an R9.4 flow cell and sequenced on a Minion device with ONT MinKNOW software.

###### England

Nanopore sequencing libraries were prepared using the 1D genomic kit (SQK-LSK108) with native barcoding kit (EXP-NBD103), according to the manufacturer's protocol. End-repaired, barcoded DNA ($\sim$ 8kb peak fragment lengths) from 3 to 7 samples were pooled into a single sequencing library and sequenced on a flow cell version R9.4.1 on either MinION or GridION.

###### South Africa

Remnant stored DNA used for Illumina WGS from each isolate was retrieved from storage and used for Nanopore library preparation. Per isolate, one microgram of undigested DNA was prepared for Nanopore sequencing using the ligation sequencing kit (SQK-LSK109). In addition, the native barcoding expansion kit (EXP-NBD104) was used for multiplexing. The protocols for sequencing genomic DNA by ligation and native barcoding were carried out according to the manufacturers' instructions. Multiplexed sequencing libraries consisted of 6-12 barcoded DNA samples, and all libraries were sequenced using SpotON R9.4.1 flow cells on a MinION device.

##### S2.4 PacBio

Thirty-five of the Malagasy samples were sequenced and processed at the Next Generation Genomics Core within Cold Spring Harbor Laboratory. Samples were quantified with a Qubit dsDNA HS Assay Kit and QC'd through a Pulsed Field Gel Electrophoresis system. Then, samples were sheared at 10kb using a Megaruptor device and size-selected to 8-10 kb with a Blue pippin instrument - followed by 0.45X ampure bead purification. The PacBio library protocol SMRTbell Express Template Prep Kit 2.0 was used for each sample. Briefly, the first step was the removal of single-stranded overhangs followed by DNA Damage Repair, End-Repair/A-tailing, Ligation of overhang barcoded adaptors and sample pooling. A total of 3 pools were produced: LID50532 (16 samples), LID50533 (10 samples), and LID50534 (9 samples). After pooling, 0.5X ampure bead clean up was performed. A Sequel I instrument was used to sequence the 3 library pools. Libraries were annealed for an hour and bounded for an hour using sequel binding kit 3.0. Bound SMRTbell complexes were then purified with ampure beads. The run was set up as 10kb length for 10 hours movie time. The Sequel 1M V2 SMRT cells were used for each library.

The circular consensus was called via the SMRTlink graphical user interface version 6.0.0.47841.

### S3 Sequencing data preparation and quality control

#### S3.1 Overview

Nanopore data were basecalled and de-multiplexed using the ONT software program Guppy (version 5.0.16). As a quality control, we removed non-*M. tuberculosis* sequencing reads by aligning all reads to a database of common contaminants[4]. Decontaminated data with mean read depth less than 20/30 (Illumina/Nanopore) were excluded from the study, as were isolates for which a single lineage could not be determined (as a proxy for potential contamination with a second strain).

#### S3.2 Details

All Nanopore data were basecalled and de-multiplexed with ONT's software guppy (version 5.0.16) using the "super high-accuracy model" (`dna_r9.4.1_450bps_sup_prom.cfg`), disabling quality score filtering and with barcode trimming when de-multiplexing. Sequencing reads were first decontaminated using reference genomes from a wide range of organisms, including viral, human, Mycobacterial and nasopharyngeal-associated bacteria as described previously[4]. Then, any reads that aligned to the *M. tuberculosis* reference genome (accession: NC_000962.3; H37Rv) were retained. For Illumina, read-pairs with only one of the pair mapped to the reference were retained. All read mapping was performed using `minimap2` (v2.17)[5] for Nanopore and `bwa mem` (v0.7.17)[6] for Illumina. Isolates with decontaminated depth less than 30x (Nanopore) or 20x (Illumina) were removed from subsequent analyses. All remaining decontaminated fastq files were randomly subsampled to a depth of 150x (Nanopore) and 60x (Illumina) using `rasusa` (v0.6.0)[7]. Any isolate with depth below this maximum threshold was left unchanged. In the last step of quality control (QC), lineages were assigned for each isolate. A panel of lineage-defining SNPs was used in conjunction with an isolate's Illumina variant calls for the lineage assignment as previously reported by[8]. If a lineage could not be determined for an isolate, or if there were multiple (major) lineages identified (indicating mixture), the isolate was not included in the analysis.

### S4 Variant calling

#### S4.1 Illumina

Illumina variant calls were made using the COMPASS pipeline[9](`https://github.com/oxfordmmm/CompassCompact`) used by the United Kingdom Health Security Agency (UKHSA)[10]. Briefly, reads were mapped to H37Rv (accession: NC_000962.3), and `samtools mpileup` (v0.1.19) was used to identify SNPs[11]. SNPs were filtered based on the following criteria: i) must have at least five high-quality (> 25) supporting bases, ii) must have at least one read in each direction, iii) 75% of reads must be high-quality, iv) the diploid genotype must be homozygous, v) fraction of reads supporting the major allele must be at least 90%. In addition, any SNPs falling within a predefined masked region, defined by aligning H37Rv to itself and identifying repetitive regions[12], were excluded.

##### S4.2 Nanopore

Nanopore reads were aligned to H37Rv using `minimap2` (v2.17), with options to produce SAM output and no secondary alignments (`-a --secondary=no`). The subsequent SAM file was provided as input to the BCFtools (v1.13;[13]) subcommand `mpileup` with the following options: ignore read-pair overlaps (`-x`), do not perform insertion and deletion (indel) calling (`-I`), minimum base quality of 13 (`-Q 13`), a homopolymer error coefficient of 100 (`-h 100`), and a maximum read length of 10,000 for the BAQ algorithm (`-M 10000`). The resulting pileup was then used to call SNPs with `bcftools call` using the multi-allelic caller with a haploid model and an option to skip indel variants (`-m --ploidy 1 -V indels`).

Only SNPs passing the following criteria were kept: i) a quality score of at least 85, ii) each strand must have at least 1% of read depth, iii) read depth at least 20% of the isolate median read depth, iv) a variant distance bias (VDB; a measure of whether a variant's position is randomly distributed within the reads supporting it) of at least 0·00001, v) fraction of reads supporting the called allele of 90% or more. SNPs were also masked in the same manner as Illumina data outlined above (Section S4.1).

##### S4.3 Pairwise SNP distances

To determine the distance between isolates, we first generated isolate consensus sequences for both Nanopore and Illumina sequencing modalities. This consensus sequence is obtained by applying the SNP calls (see Variant calling) to the *M. tuberculosis* reference genome H37Rv. We exclude any positions where i) the position failed filtering, ii) the genotype is null, or iii) the position is within the reference mask (see Section S4.1). A pairwise distance matrix was calculated using `psdm` (v0.1.0)[14].

#### S5 Consensus genomic sequence assembly

The eight PacBio isolates with read depth over 30x were assembled using Flye (v2.8)[15] with one polishing iteration and input type `--pacbio-hifi`. Assembly contigs were removed if they were classified by Centrifuge[16] as not being part of the *M. tuberculosis* Complex (MTBC; taxon ID: 77643). During this contig decontamination process, one isolate was found to have chromosomes for three different species, and the PacBio data was therefore discarded leaving us with seven PacBio assemblies. We built a gold-standard reference sequence for each isolate using the unpolished PacBio assembly along with a mask for low-quality regions identified by aligning the isolates' Illumina reads to the PacBio assembly and flagging any position with either less than 10 reads mapping to it or less than 90% agreement. These assemblies are used as a "truth" sequence for the respective isolates when assessing SNP precision and recall.

##### S5.1 Evaluation of precision/recall using PacBio assemblies as truth

Evaluation of precision/recall was done with `varifier` (v0.3.1)[17] to generate a list of true (expected) variants between the isolate's assembly and the *M. tuberculosis* reference sequence, and using the tool `hap.py` (v0.3.14)[18] to evaluate precision and recall.

#### S6 Extended clustering metrics definitions

##### S6.1 Outline

We treat Illumina as the established standard when comparing clustering. We define the Illumina clustering as I and the Nanopore clustering as N. To quantify the recall and precision of the Nanopore clustering, we compared the clustering graphs I and N with three similarity metrics.

We define the sample-averaged cluster recall (SACR) by calculating, for each clustered isolate, $s$, in $I$, the proportion of isolates in its Illumina cluster ($C_{s,I}$) also present in its Nanopore cluster ($C_{s,N}$), and then averaging this over all isolates. We likewise define the sample-averaged cluster precision (SACP) as the proportion of isolates in $C_{s,N}$ also present in $C_{s,I}$ averaged over all isolates. SACR indicates whether isolates have been missed by Nanopore clustering (false negatives), and SACP reflects additional isolates being clustered by Nanopore (false positives).

One shortcoming of SACR and SACP is that they do not account for Nanopore clusters composed solely of Illumina singletons. Therefore, we define the excess clustering rate (XCR) as the proportion of Illumina singletons that are clustered by Nanopore. A value of 0·1 would indicate that 10 percent of Illumina singletons were part of a Nanopore cluster.

##### S6.2 Details

As outlined above, to assess how closely Nanopore SNP-based clustering approximates Illumina SNP-based clustering, we adapt a similarity measure on sets; the Tversky Index[19]. We define the Illumina clustering as $I$ and the Nanopore clustering as $N$. We are interested in being able to quantify the recall and precision of the Nanopore clustering with respect to Illumina. In this sense, recall describes the proportion of clustered samples (isolates) in $I$ clustered with the expected (correct) samples in $N$. Likewise, precision in this context tells us when extra samples are added to existing clusters by $N$ or when clusters in $I$ are joined in $N$.

In order to be able to define precision and recall when comparing two clustering graphs $I$ and $N$, we define the Tversky Index

$$TI(n, I, N) = \frac{|C_{n,I} \cap C_{n,N}|}{|C_{n,I} \cap C_{n,N}| + \alpha|C_{n,I} - C_{n,N}| + \beta|C_{n,N} - C_{n,I}|} \tag{S1}$$

where $C_{n,I}$ is the cluster in $I$ that sample $n$ is a member of. When $\alpha = 1$ and $\beta = 0$ in Equation S1, we get a metric analogous to recall - as described above. Therefore, we define recall, $R$, for a single sample $n$ as

$$R(n, I, N) = \frac{|C_{n,I} \cap C_{n,N}|}{|C_{n,I} \cap C_{n,N}| + |C_{n,I} - C_{n,N}|} = \frac{|C_{n,I} \cap C_{n,N}|}{|C_{n,I}|} \tag{S2}$$

When $\alpha = 0$ and $\beta = 1$ in Equation S1, we get a metric analogous to precision. As such, we define precision $P$, for a single sample $n$ as

$$P(n, I, N) = \frac{|C_{n,I} \cap C_{n,N}|}{|C_{n,I} \cap C_{n,N}| + |C_{n,N} - C_{n,I}|} = \frac{|C_{n,I} \cap C_{n,N}|}{|C_{n,N}|} \tag{S3}$$

With these definitions for a single sample, we can assess the recall and precision of the Nanopore clustering, $N$, with respect to the Illumina clustering, $I$, by averaging each metric over all samples in $I$. This gives us the *Sample-Averaged Cluster Recall* (SACR)

$$SACR = \frac{\sum_{n}^{V_I} R(n, I, N)}{|V_I|} \tag{S4}$$

where $V_I$ is the set of samples (nodes) in $I$ (Illumina graph). Likewise, we define the *Sample-Averaged Cluster Precision* (SACP) as

$$SACP = \frac{\sum_{n}^{V_I} P(n, I, N)}{|V_I|} \tag{S5}$$

SACR states, on average, what proportion of the samples clustered together in $I$ are also clustered together in $N$ (Nanopore) - it is a measure of how many true positives Nanopore retains. Inversely, SACP states, on average, what proportion of the samples clustered together in $N$ are also clustered together in $I$ - it is a measure of how many extra samples Nanopore adds to clusters.

However, SACR and SACP do not inherently account for when $N$ has clusters containing only samples deemed non-clustered (singleton) in $I$. In order to quantify any extra clustering by $N$, we establish the *Excess Clustering Rate* (XCR) as the proportion of singletons (disconnected nodes) in $I$ that are connected in $N$. We define XCR as

$$XCR = \frac{|S_I - S_N|}{|S_I|} \tag{S6}$$

where $S_I$ and $S_N$ are the sets of singletons in the respective graphs.
We assess the cluster similarities using the Python programming language with the `networkx` library[20]. For a given threshold, we create the Illumina clustering (graph), $I$, and the Nanopore clustering, $N$ - from the relevant distance matrix - and use these to calculate the SACR, SACP, and XCR using Equation S4, Equation S5, and Equation S6, respectively.

### S7 Illustrated example of clustering metrics

Section S6 outlines three metrics - SACR, SACP and XCR - for evaluating the similarity between two different strategies for transmission clustering. In order to provide the reader with greater intuition for the purpose of each metric, we present an illustrated example in Figure S1.

We take Figure S1a to be the truth clusters and Figure S1b to be test clusters. These are akin to Illumina ($I$) and Nanopore clusters ($N$), respectively, in Section S6. The individual recall and precision values (defined in Equation S2 and Equation S3) for each sample in Figure S1a are shown in Table S1. SACR and SACP (defined in Equation S4 and Equation S5) are *sample-averaged*, so their values for this example are 0·82 and 0·83 respectively.

To highlight the objective of SACR, we use the truth and test clusters containing the sample $F$. Samples $F$, $G$, $H$ and $I$ are shared between both, but $J$ is missing from the test cluster. To calculate the individual recall for $F$, we take the intersection size of the truth and test clusters it exists in and divide it by the size of the truth cluster - $\frac{4}{5} = 0{\cdot}8$. We do the same for the precision of sample $D$, except we divide by the size of the test cluster - giving $\frac{2}{3} = 0{\cdot}66$.

The relevance of the XCR metric is best exemplified by the test cluster containing samples $L$ and $M$. As we calculate SACR and SACP for all samples in the *truth* clusters, these two samples would be ignored. However, they are samples that - according to the truth - should not be part of any cluster (singletons). Therefore, SACR and SACP cannot capture these extra clusterings if they do not contain clustered truth samples. XCR covers this limitation and is the proportion of singletons in the truth that are clustered in the test (see Equation S6). As Figure S1 does not show singletons, let us pretend there are 20 singletons in the truth (including samples $L$ and $M$). This would give an XCR of $2/20 = 0{\cdot}1$.

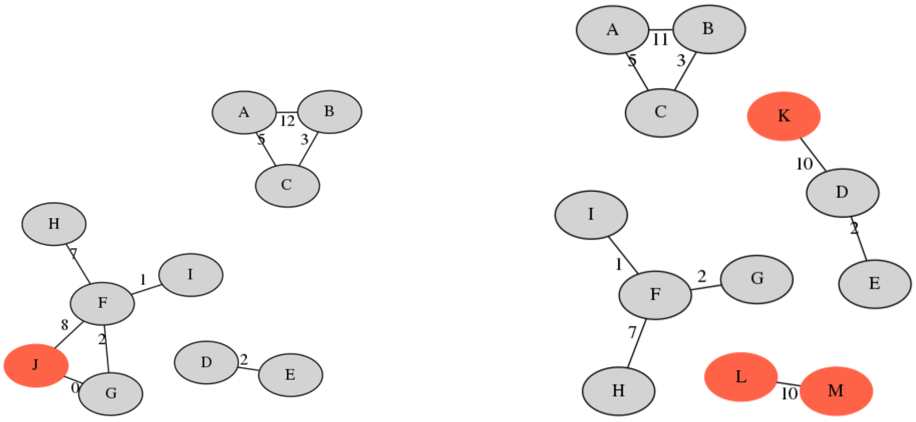


**(a)** Truth clusters

**(b)** Test clusters

**Figure S1:** Illustrative examples of transmission clustering. **a)** represents truth clusters, while **b)** is clustering from some “test” method we would like to compare to **a**. The nodes represent samples with the numbers on the edges connecting them indicating the distance between those two samples. The red nodes indicate samples with a clustering disparity between the two clusterings. Note, we do not show singletons (disconnected nodes) - e.g., *J* is missing from **(b)**.

| sample | recall | precision |
|---|---|---|
| A | 1·0 | 1·0 |
| B | 1·0 | 1·0 |
| C | 1·0 | 1·0 |
| D | 1·0 | 0·66 |
| E | 1·0 | 0·66 |
| F | 0·8 | 1·0 |
| G | 0·8 | 1·0 |
| H | 0·8 | 1·0 |
| I | 0·8 | 1·0 |
| J | 0·0 | 0·0 |

**Table S1:** Cluster recall and precision results for each sample in Figure S1.

### S8 Drug susceptibility testing methods

#### S8.1 Madagascar

Culture on Löwenstein-Jensen (LJ) is still the gold-standard method for *M. tuberculosis* identification and the detection of resistance. The indirect proportion method on LJ medium was performed to test the susceptibility of positive cultures against anti-*M. tuberculosis* drugs. $4\,\mu g\,mL^{-1}$, $0.2\,\mu g\,mL^{-1}$, $40\,\mu g\,mL^{-1}$, $2\,\mu g\,mL^{-1}$, $30\,\mu g\,mL^{-1}$, $30\,\mu g\,mL^{-1}$, and $40\,\mu g\,mL^{-1}$ were the critical concentrations used for Streptomycin, Isoniazid, Rifampicin, Ethambutol, Kanamycin,

Amikacin and Capreomycin, respectively. The growth on a drug-free medium was compared with the growth on a medium containing an anti-*M. tuberculosis* agent. An isolate was identified as resistant if at least 1% of growth is present at the critical concentration of the drug in the culture medium.

##### S8.2 South Africa

Isoniazid, ofloxacin, amikacin, and ethambutol (two concentrations) phenotypic DST was performed on Middlebrook 7H with critical concentrations $0.2\,\mu g\,mL^{-1}$, $2.0\,\mu g\,mL^{-1}$, $4.0\,\mu g\,mL^{-1}$, and $7.5\,\mu g\,mL^{-1}$ and $10.0\,\mu g\,mL^{-1}$, respectively.

#### S9 Drug resistance predictions with Mykrobe

We used Mykrobe (v0.10.0)[21] to obtain predictions of each isolates' drug susceptibility profile for 11 drugs - using the `predict` subcommand. For both technologies, we used a haploid model (`--ploidy haploid`) and a proportion of expected depth of 20% (`--min_proportion_expected_depth 0.2`). We set the expected error rate to 0·001 (`-e 0.001`) for Illumina and 0·08 (`-e 0.08`) for Nanopore. (We did not use the preset Nanopore settings (`--ont`).)

With Illumina data, Mykrobe can improve resistance predictions by detecting minor (low frequency, within-isolate) alleles, although this is not standard practice in the community. Including minor alleles resulted in elevated levels of discordance between Nanopore and Illumina, as many insertions and deletions were only (erroneously) detected with the Nanopore data. We therefore used a haploid model for both sequencing modalities.

##### S9.1 Mutation concordance

We assessed the concordance of Mykrobe's Nanopore and Illumina genotypes for each of the 66,537 nucleotide-level mutations. In order to get genotypic information on all mutations in Mykrobe's catalogue, we ran Mykrobe with the `-A` option and `--format json` to output the results in the JSON format. For a given isolate, we compare the genotype of each mutation in both the Illumina and Nanopore JSON file. If there are any filters present (e.g., low coverage) for the mutation in either technology's output, we skip the mutation; otherwise, we consider the mutation concordant if the genotype is exactly the same between the two technologies.

In total, we found four genotype discordances. Three of these discrepant mutations were *katG* 1bp deletions at consecutive positions within a homopolymer in *katG*, all in the same isolate, effectively describing one deletion event - thus only affecting a single phenotype call. The other discrepancy was a *katG* 1bp deletion in a separate isolate.

We then look at how these genotype differences translate into different predictions, which can differ subtly when there are no-calls. There were five false positive (FP) resistance calls made by Mykrobe with Nanopore data, with respect to the Illumina predictions. Three of the FPs (amikacin, capreomycin, and kanamycin) are caused by one mutation (*rrs* a1401g) in a single isolate, which confers resistance to multiple drugs. There was also a discordant streptomycin call, due to *rrs* a514c. In both of these *rrs* FPs, the mutation was in fact detected by Illumina, but was filtered out due to low coverage (17 percent and 19 percent of expected coverage, respectively); the Nanopore data had very good depth for both of these mutations. This was not counted in the discordance count in the previous paragraph, as that counted variants where Illumina/Nanopore made actively conflicting genotype calls, whereas here there was no Illumina call (the call was filtered). There was also one isoniazid discordance, due to a frameshift deletion in *katG* (mentioned above) erroneously called by Nanopore. When consulting the DST phenotype for these discrepancies, Nanopore had the correct predictions for the two *rrs* mutations, and Illumina had the correct prediction for the *katG* deletion. See Table S3 for a full description of the Mykrobe genotypic and phenotype prediction discrepancies.

#### S10 Full dataset distance plot

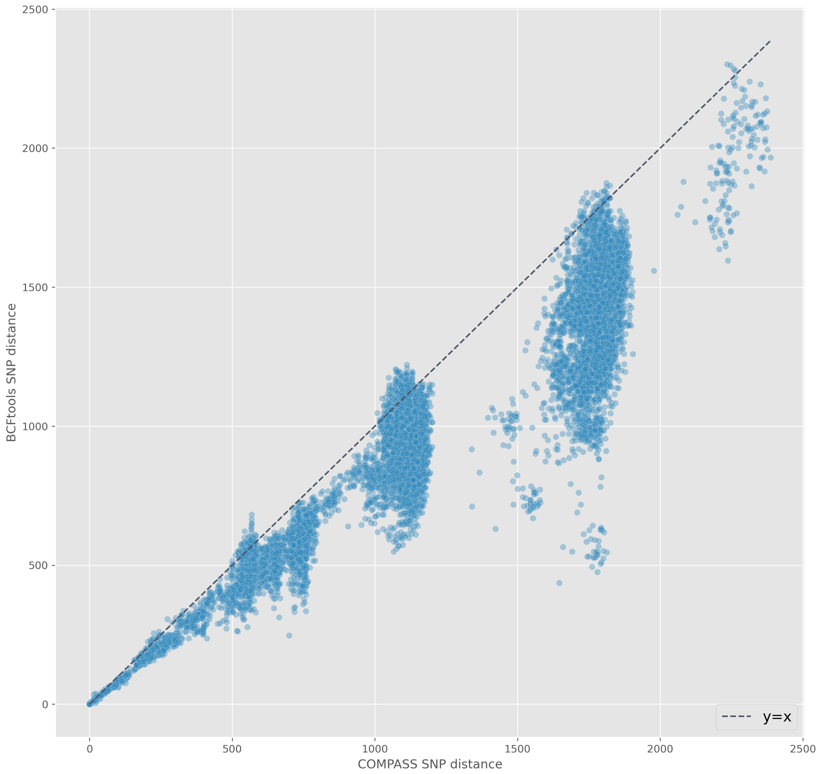


**Figure S2:** Pairwise SNP distance relationship between Illumina (COMPASS; x-axis) and Nanopore (`bcftools`; y-axis). Each point represents the SNP distance between two isolates. The black, dashed line shows the identity line (i.e. $y = x$).

#### S11 Selecting Nanopore SNP distance thresholds

In order to select the Nanopore SNP distances that provide the most similar clustering to Illumina, we calculate the SACR, SACP, and XCR for a range of values (2-14). For the two Illumina distance thresholds 5 and 12, we create the Illumina clusters for the relevant distance threshold and compare those to the Nanopore clustering for each of values between 2 and 14. We then select the Nanopore threshold that provides the best balance of SACR, SAP, and XCR for the relevant Illumina threshold. Figure S3 shows the results of these threshold evaluations. Based on these, we select Nanopore distance thresholds 6 and 12 to correspond to Illumina thresholds 5 and 12, respectively.

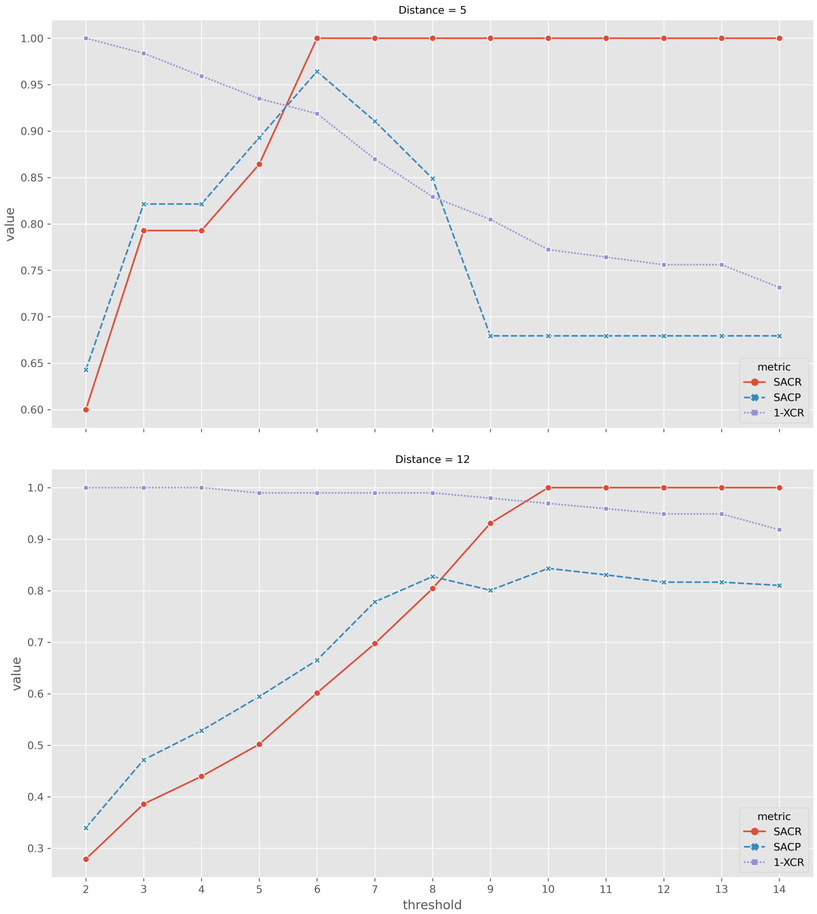


**Figure S3:** Illumina and Nanopore (`bcftools`) transmission cluster similarity for various SNP distance threshold. Each subplot compares the Nanopore clustering for the threshold on the x-axis to the Illumina clustering based on the distance (threshold) in the subplot title. SACR (red), SACP (blue), and 1−XCR are represented by the solid, dashed, and dotted lines, respectively. SACR=sample-averaged cluster recall; SACP=sample-averaged cluster precision; XCR=excess clustering rate.

### S12 Mixed modality self-distance

The "self-distance" for each isolate is the distance between an isolate's Illumina and Nanopore data. As the sequencing data in this study originate from the same source, we know the self-distance for any isolate *should* be 0. However, we also know there are major technical differences between Illumina and Nanopore; therefore, small variability in self-distance is likely. We plot the self-distances in Figure S4 and see that 66% (100/151) of the isolates have a distance of 0 between their Illumina (COMPASS) and Nanopore (`bcftools`) data, with 86% (130/151) less than 3 SNPs apart. All isolates have a self-distance of less than 10, except one isolate (`mada_1-33`), which has a self-distance of 42. We investigated the possibility of a sample mix-up being the cause of this discrepancy but could not find any such convincing evidence.

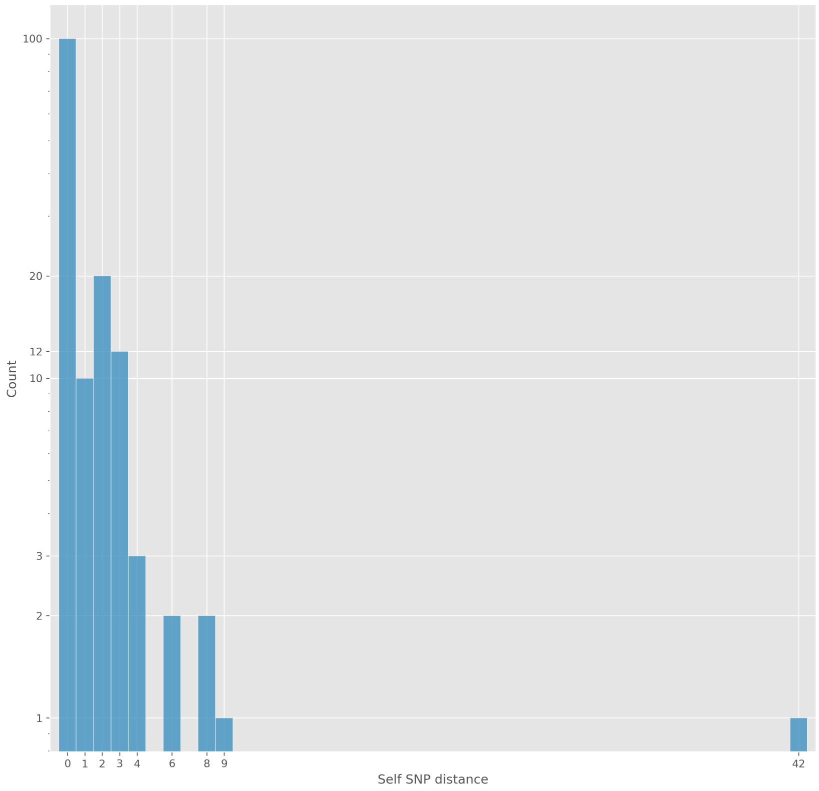


**Figure S4:** Mixed modality "self-distance". This plot shows the SNP distance (x-axis) between each isolate's COMPASS (Illumina) and `bcftools` (Nanopore) VCF calls. Note, the y-axis is log-scaled.

#### S13 Mixed sequencing modality distance plot

The outliers in the inset window of Figure S5 are due to discrepant SNP calls between some isolate's Illumina and Nanopore data in the gene *ppe54*. There are some gaps in the genome mask in this gene, and nearly all of the discrepancies lie within one of these gaps.

In general, what is happening is one isolate's Illumina calls good quality SNPs (or reference alleles) in these regions, but the same isolate's Nanopore has filtered calls for low quality and/or low fraction of read support for those sites. Then, the inverse happens in the other isolate - i.e., the Illumina generally has no coverage in the gaps but the Nanopore has good quality calls. So, when you do the same-technology distances (Figure 3 in the main text), these sites are all ignored due to filtering on one of the sequencing modalities, but when we assessing mixed-modality distance, these sites lead to larger-than-expected distances.

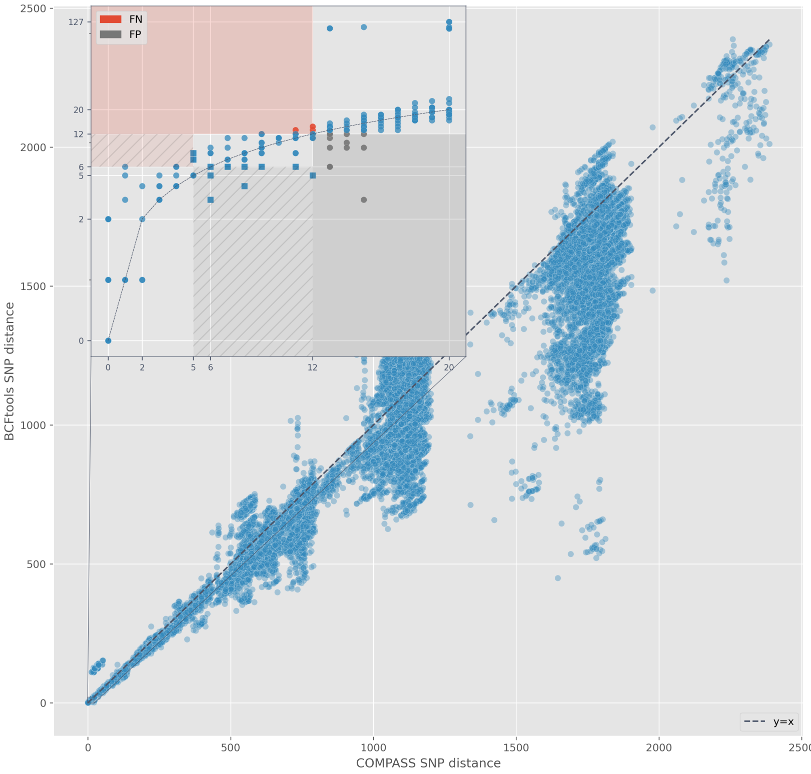


**Figure S5:** Pairwise SNP distance relationship between Illumina (COMPASS; x-axis) and mixed COMPASS-`bcftools` calls (y-axis). Each point represents the SNP distance between two isolates. The black, dashed line shows the identity line (i.e., y=x). The zoomed inset shows all pairs where the COMPASS distance is $\leq 20$. The red area and points indicate pairs with a mixed distance $\geq 12$, but an Illumina distance $\leq 12$. These pairs are deemed false negative (FN) connections. The red area with stripes indicates pairs that are FN connections at an Illumina threshold of 5 (mixed threshold 6), but not when the threshold is expanded to 12. These pairs are shown as square points. The grey area and points is the inverse - i.e., false positive (FP) connections. Note, the y-axis in the inset window is log-scaled.

#### S14 Effect of Nanopore read depth on drug resistance predictions

An important consideration when using Nanopore sequencing data for DST prediction is how much data is needed. The quantity of data required has implications for how long the Nanopore sequencing device needs to be run or how many samples can be multiplexed in a single run in order to yield sufficient data for reliable predictions. A previous study by Votintseva *et al.* found that deep coverage is required of Nanopore to predict drug resistance accurately[22].

As our dataset contains a broad range of Nanopore read depths (29-150x), we explore whether this requirement of high coverage still holds. As Figure S6 illustrates, there is no relationship between Nanopore read depth and erroneous predictions. If low read depth lead to poor resistance prediction, we would expect the proportions of FPs (red) and FNs (purple) in the low-depth bins to be greater than in the high-depth bins - which is not the case.

Given these results, we found no clear evidence that depth as low as 30x leads to an increased level of false phenotype predictions. To put this low-depth result into perspective, we can do a "back of the envelope" calculation to determine how long a Nanopore device needs to be operated to achieve 30x coverage theoretically. First, we use a conservative yield from Smith *et al.* of 11 gigabases (total) per 24 hours with six samples multiplexed on a single flow cell[23]. Assuming equal depth from each sample, this equates to 76·4Mb/hour/sample - dividing by the *M. tuberculosis* genome size (4·41Mb) gives an hourly coverage yield of 17·3x per sample. Finally, we divide our desired yield, 30x, by this per-sample hourly yield to get a required device runtime of 1 hour and 45 minutes. In contrast, to get 100x of data (the target coverage in[23]), the Nanopore device would need to be run for 5 hours and 47 minutes. Thus, in theory, one can save 4 hours of sequencing, use more samples per flow cell, decrease costs as a result, and still obtain reliable drug resistance predictions.

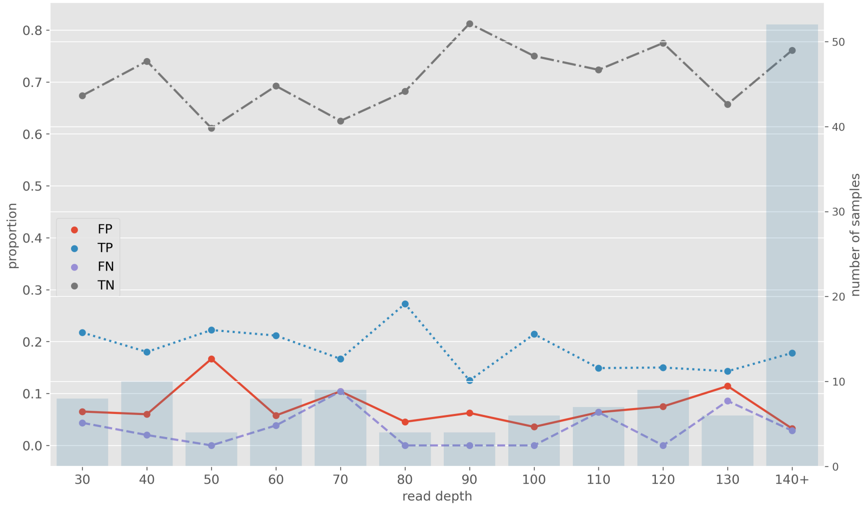


**(a)**

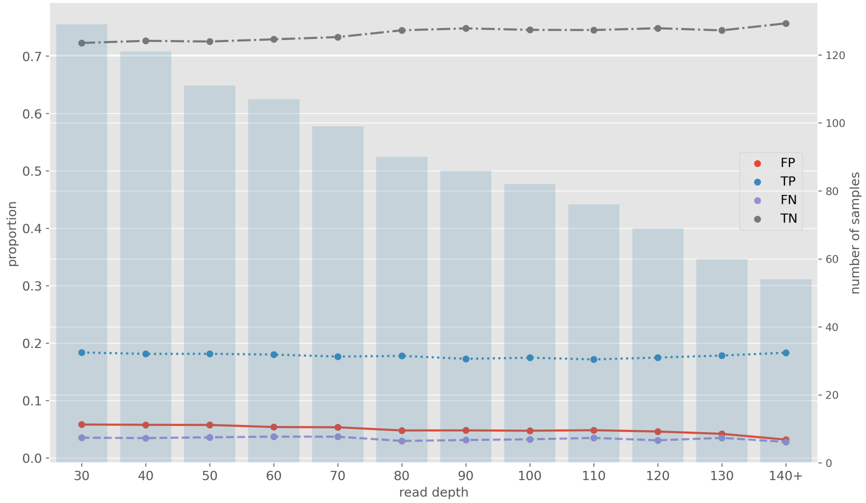


**(b)**

**Figure S6:** Effect of Nanopore read depth on Mykrobe DST prediction. Each point indicates the proportion (left y-axis) of classifications of that type at the read depth on the x-axis. The blue bars indicate the number of samples (right y-axis) contained in each bin. **(a)** read depth, $d$, is binned such that 40 is all samples where $40 \leq d \leq 50$. **(b)** read depth bin 40 is all samples where $d \geq 40$. FP=false positive (red); TP=true positive (blue); FN=false negative (purple); TN=true negative (grey).

#### S15 Genotypic drug resistance prediction concordance with phenotype

| Drug | Technology | FN(R) | FP(S) | FNR(95% CI) | FPR(95% CI) | PPV(95% CI) | NPV(95% CI) |
|---|---|---|---|---|---|---|---|
| Isoniazid | Illumina | 9(51) | 3(48) | 17·6% (9·6-30·3%) | 6·2% (2·1-16·8%) | 93·3% (82·1-97·7%) | 83·3% (71·3-91·0%) |
| Isoniazid | Nanopore | 9(51) | 4(48) | 17·6% (9·6-30·3%) | 8·3% (3·3-19·6%) | 91·3% (79·7-96·6%) | 83·0% (70·8-90·8%) |
| Rifampicin | Illumina | 6(48) | 1(44) | 12·5% (5·9-24·7%) | 2·3% (0·4-11·8%) | 97·7% (87·9-99·6%) | 87·8% (75·8-94·3%) |
| Rifampicin | Nanopore | 6(48) | 1(44) | 12·5% (5·9-24·7%) | 2·3% (0·4-11·8%) | 97·7% (87·9-99·6%) | 87·8% (75·8-94·3%) |
| Ethambutol | Illumina | 4(14) | 14(77) | 28·6% (11·7-54·6%) | 18·2% (11·2-28·2%) | 41·7% (24·5-61·2%) | 94·0% (85·6-97·7%) |
| Ethambutol | Nanopore | 4(14) | 14(77) | 28·6% (11·7-54·6%) | 18·2% (11·2-28·2%) | 41·7% (24·5-61·2%) | 94·0% (85·6-97·7%) |
| Streptomycin | Illumina | 4(8) | 11(83) | 50·0% (21·5-78·5%) | 13·3% (7·6-22·2%) | 26·7% (10·9-52·0%) | 94·7% (87·2-97·9%) |
| Streptomycin | Nanopore | 3(8) | 11(83) | 37·5% (13·7-69·4%) | 13·3% (7·6-22·2%) | 31·2% (14·2-55·6%) | 96·0% (88·9-98·6%) |
| Amikacin | Illumina | 1(11) | 2(78) | 9·1% (1·6-37·7%) | 2·6% (0·7-8·9%) | 83·3% (55·2-95·3%) | 98·7% (93·0-99·8%) |
| Amikacin | Nanopore | 0(11) | 2(78) | 0·0% (0·0-25·9%) | 2·6% (0·7-8·9%) | 84·6% (57·8-95·7%) | 100·0% (95·2-100·0%) |
| Capreomycin | Illumina | 1(1) | 1(51) | 100·0% (20·7-100·0%) | 2·0% (0·3-10·3%) | 0·0% (0·0-79·3%) | 98·0% (89·7-99·7%) |
| Capreomycin | Nanopore | 1(1) | 1(51) | 100·0% (20·7-100·0%) | 2·0% (0·3-10·3%) | 0·0% (0·0-79·3%) | 98·0% (89·7-99·7%) |
| Kanamycin | Illumina | 0(0) | 1(52) | - | 1·9% (0·3-10·1%) | 0·0% (0·0-79·3%) | 100·0% (93·0-100·0%) |
| Kanamycin | Nanopore | 0(0) | 1(52) | - | 1·9% (0·3-10·1%) | 0·0% (0·0-79·3%) | 100·0% (93·0-100·0%) |
| Ofloxacin | Illumina | 0(10) | 4(77) | 0·0% (-0·0-27·8%) | 5·2% (2·0-12·6%) | 71·4% (45·4-88·3%) | 100·0% (95·0-100·0%) |
| Ofloxacin | Nanopore | 0(10) | 4(77) | 0·0% (-0·0-27·8%) | 5·2% (2·0-12·6%) | 71·4% (45·4-88·3%) | 100·0% (95·0-100·0%) |

**Table S2:** Comparison of WGS-based drug resistance predictions with culture-based phenotypes. For this comparison, we use phenotypic DST as the reference standard and evaluate Mykrobe Illumina and Nanopore resistance predictions accordingly. FN=false negative, meaning Nanopore does not detect resistance where Illumina does; R=number of resistant isolates; FP=false positive, meaning Nanopore detects resistance where Illumina finds susceptible; S=number of (Illumina) susceptible isolates; FNR=false negative rate; FPR=false positive rate; PPV=positive predictive value; NPV=negative predictive value; CI=Wilson score confidence interval.

| Isolate | Mutation | Genotype | Genotype discordant | Phenotype (predicted) | Phenotype discordant |
|---|---|---|---|---|---|
| `mada_135` | *katG* gc1037c | 0 / 1 | YES | S / R | YES |
| `mada_135` | *katG* cc1038c | 0 / 1 | YES | S / R | YES |
| `mada_135` | *katG* cc1039c | 0 / 1 | YES | S / R | YES |
| `mada_1-41` | *katG* cc1038c | 0 / 1 | YES | R* / R | NO* |
| `R26791` | *rrs* a1401g | F / 1 | NO† | S / R | YES |
| `mada_1-3` | *rrs* a514c | F / 1 | NO† | S / R | YES |

**Table S3:** Discordances between Mykrobe genotype calls, and Mykrobe predicted phenotype when comparing Illumina and Nanopore. In the genotype column the Illumina and Nanopore genotype calls are shown separated by a forward-slash Illumina/Nanopore. F = call was filtered out; 0 = reference allele; 1 = alternate allele. The "Genotype discordant" column shows whether this difference was considered a discordance in our calculation. The "Phenotypes (predicted)" column shows the Mykrobe predictions for Illumina/Nanopore: S = susceptible; R = resistant. The "Phenotype discordant" column shows whether the predicted phenotypes in the previous column are the same.

*The isolate had other mutations in *katG* that cause isoniazid resistance.

†We skip mutations where either technology's call is filtered (see Section S9.1).
